# Interactive Effects of Telomere Length and Genetic Variants on Alzheimer Disease Risk Across Multiple Ancestral Populations

**DOI:** 10.64898/2025.12.15.25342309

**Authors:** Zainab Khurshid, Tong Tong, Oluwatosin Olayinka, John J. Farrell, Congcong Zhu, Alzheimer’s Disease Sequencing Project, Eden R. Martin, Will Bush, Margaret A. Pericak-Vance, Li-San Wang, Gerard C. Schellenberg, Jonathan L. Haines, Kathryn L. Lunetta, Xiaoling Zhang, Lindsay A. Farrer

## Abstract

**Background:** Telomere length (TL), a biomarker of biological aging, but its association with Alzheimer’s disease (AD) remains unclear.

**Methods:** We estimated TL in whole-genome sequencing data from 35,014 Alzheimer’s Disease Sequencing Project participants using TelSeq, which after quality control yielded a dataset including 6,973 persons of European ancestry (EA), 4,188 African Americans (AA), 4,005 Caribbean Hispanics (CH), and 4,170 Native American Hispanics (NAH). TL was log-transformed, adjusted for age and blood cell counts, and z-scaled. Scaled TL was dichotomized into long and short groups according to the median. An AD GWAS for the interaction of TL with variants having a minor allele count >20 was performed in each ancestry group using logistic regression models including SNP and TL main effects and a SNP×TL interaction term.

**Results:** AD risk was associated with shorter TL (β = -0.18, *P* < 2×10^-16^). Longer TL was associated with dosages of *APOE* ε2 (*P*<5.08×10^-8^) and *APOE* ε4 (*P*=2.10×10^-2^). In the EA group, genome-wide significant (GWS) TLxSNP interactions were identified for variants in *SEMA6A (P*=1.42×10^-8^) and *LOC105378654 (P*=4.17×10^-8^), between *IL15* and *INPP4B (P*=1.77×10^-8^) and upstream of *RP11-2N5.2* (*P*=4.60×10^-8^). In the NAH group, GWS interactions were observed with an intronic variant in *BSN* (*P*=3.26×10^-8^) and missense variant in *MST1* (*P*=3.26×10^-8^). In the total sample, interactions with variants between *CTD-2160D9.1* and *EEF1A1P20* (*P*<1.19×10^-8^), in *TBC1D22A* (*P*=1.06×10^-8^) and in *PLK1* (*P*=3.28×10^-8^) were GWS.

**Conclusion:** We identified variants that significantly impact AD risk through their interaction with TL, suggesting that TL maintenance pathways may be central to AD pathogenesis.

## Background

Alzheimer disease (AD) is a neurodegenerative disease that causes cognitive decline and dementia usually after age 65. It is predicted that by 2060, the number of people in the US affected with AD will grow to 13.8 million [1]. Despite its high heritability (*h^2^*L=L0.58–0.79), the genetic basis of AD is not fully understood. In addition to the *APOE* ε4 allele and genetic variants at >75 loci identified by genome-wide association studies (GWAS) [2] which accounts for a substantial portion of the genetic variance of AD [3] there are many non-genetic factors associated with the risk of AD [4].

Telomeres are long, repeated TTAGGG sequences at the end of chromosomes which shorten with each cell cycle due to incomplete replication [5] and maintains genomic stability by preventing chromosome fusion. Telomere length (TL), a heritable biomarker of biological age [6,7,8], is also affected by environmental exposures [9] and genetically determined factors including telomerase reverse transcriptase (*TERT*) which regulates telomere maintenance.10 TL has been associated with many age-related diseases including AD [11]. TL is sensitive to oxidative stress and inflammation, and shortening has been linked to cognitive impairment, amyloid aggregation, and tau phosphorylation in AD [12]. Although many studies have examined genetic variants associated with either AD3 and TL [13], as well as their relationship [14,15,16], the role of TL in AD remained complex, potentially varying across disease stages [17]. Prior studies examining the relationship between TL and AD risk have focused on European ancestry (EA) cohorts, despite known ancestral differences in TL [18]. To address this gap and identify genetic modulators of the influence of TL on AD risk, we performed an interaction genome-wide association study (GWAS) using the multi-ancestry Alzheimer’s Disease Sequencing Project (ADSP) dataset.

## Methods

### Study participants

Study participants were selected from the ADSP whole genome sequencing (WGS) release 4 (R4) dataset (n = 35,014), processed by the Genome Center for Alzheimer’s Disease (GCAD). The pre-filtered dataset included individuals who self-identified as EA (n=15,842), Caribbean Hispanic (CH, n=5,601), South Asian (SA, n = 2,983), Hispanic (n=5,174), and African American (AA, n=5,414) (**Additional file 2: Table S1**). To more precisely reflect ancestral origins, we subsequently referred to the Hispanic individuals, most of whom were ascertained in the southwest region of the US, as Native American Hispanic (NAH), thereby distinguishing them from CH individuals. A subset of the WGS dataset contained families with two or more members affected by AD. Details of the contributing studies that provided DNA specimens and phenotype data for the ADSP participants are available at the National Institute on Aging Genetics of Alzheimer Disease Data Storage site (NIAGADS) [19].

### Genotype calling, data processing, and quality control (QC)

GCAD provided jointly called and quality-controlled (QC’ed) project-level VCF (pVCF) files via NIAGADS Data Sharing Service [19]. Details of data generation have been reported elsewhere [20,21]. Briefly, sequencing data were processed using the Variant Calling Pipeline and data management tool (VCPA), developed by GCAD for sequence data processing and joint variant calling [22]. The Genome Analysis toolkit (GATK), as implemented in VCPA, was used to generate genomic VCF (gVCF) files and pVCF files, aligned to the human GRCh38/hg38 reference genome. Variants that failed the GATK variant calling or QC protocols, were not in Hardy-Weinberg Equilibrium (P ≤ 1×10^-8^), or had a call rate <80%, mean depth > 500, allele balance for heterozygous calls (ABHet ratio) outside the range 0.30 ≤ or ≥ 0.70, PCR_P (the - log10P value of allele frequency difference between PCR-Plus and PCR-Free sequencing samples) > 7.3, or minor allele count (MAC) ≤ 20 were excluded.

We excluded WGS data derived from brain because their TL distributions were highly irregular compared to the blood derived samples, and brain DNA is more prone to technical artifacts such as contamination, which can directly bias TL estimation. PCR-amplified samples were removed because amplification distorts repeat reads and inflates TL estimates. Shorter read length samples (100/101 bp) that showed inflated TL estimates and high variance were excluded to avoid including inaccurate TL estimates. We also excluded Progressive Supranuclear Palsy (PSP) and Corticobasal Degeneration (CBD) cases, family-based study cohorts due to inflated TL estimates with much higher variance among ancestries as compared to case-control study samples, and the SA cohort due to the small proportion of cases (n=224) who were classified as undifferentiated dementia on the basis of Clinical Dementia Rating (CDR) [23]. In addition, duplicate samples and samples with missing phenotypes (AD status, age, and sex) were removed. After QC, a total of 19,336 blood samples remained (**Additional File 1: Fig. S1**), including 6,973 EA (3,701 cases/3,273 controls), 4,005 CH (1,587 cases/2,418 controls), 4,170 NAH (754 cases/3,416 controls) and 4,188 AA (1,329 cases/2,859 controls) (**Table 1**).

**Table 1.** Subject characteristics.

| AD cases (N =7371) |  |  |  |  |
| --- | --- | --- | --- | --- |
| Ancestry | N | Mean Age (SD) | % Female | APOE ε4 carrier (%) |
| EA | 3701 | 69.91(10.61) | 52.93% | 60.04% |
| CH | 1587 | 76.14(8.04) | 68.81% | 37.68% |
| NAH | 754 | 73.01(9.35) | 62.47% | 31.83% |
| AA | 1329 | 75.05(8.62) | 72.38% | 61.10% |
| controls (N=11965) |  |  |  |  |
| Ancestry | N | Mean Age (SD) | % Female | APOE ε4 carrier (%) |
| EA | 3272 | 79.14 (8.05) | 62.47% | 28.80% |
| CH | 2418 | 74.6(7.31) | 69.30% | 23.78% |
| NAH | 3416 | 67.41(9.88) | 62.82% | 18.73% |
| AA | 2859 | 73.99(7.98) | 72.71% | 36.73% |

AD = Alzheimer disease; EA = European ancestry; CH = Caribbean Hispanic; NAH = Native American Hispanic; AA = African American

### Genetic Relationship and Population Substructure

Variants with minor allele frequency (MAF) ≥ 0.05, call rate > 95%, and pairwise linkage disequilibrium (LD) r^2^ < 0.1 within a 1,000 SNP window (sliding by 50 SNPs) were retained for principal component (PC) analysis of population substructure and genetic relationship matrix (GRM) calculation using PLINK and the GENetic EStimation and Inference in Structured samples (GENESIS) R package [24,25]. The GENESIS PC-AiR method, which infers robust population structure in the presence of known or cryptic relatedness, was employed to calculate PCs for unrelated individuals and which were then projected onto the remaining related individuals. Individuals were clustered using a Gaussian Mixture Model [26] applied to multiple K-means solutions and subsequently assigned to clusters that most closely align with EA (3,701 AD cases, 3,273 controls), CH (1,587 AD cases and 2,418 controls), NAH (754 AD cases and 3,416 controls), and AA (1,329 AD cases and 2,859 controls) group]. Their characteristics are summarized in **Table 1**. As expected, individuals in the CH cluster showed greater African ancestry proportions than those in the NAH cluster (**Additional File 1: Fig. S2)**. PCs were then recalculated within each of the four ancestry clusters.

### Telomere Length Estimation and Polygenic Risk Score Calculation

The WGS CRAM files were processed using TelSeq to estimate TL [27] Reads containing more than seven consecutive TTAGGG motifs were defined as telomeric. Blood-derived TL has been shown to be a reliable proxy overall for most tissue types [6]. TL values were log-transformed and adjusted for chronological age and blood cell counts to mitigate confounding effect [28]. We used polygenic risk scores (PRS) as proxies for blood cell counts to account for variation in TL due to blood cell type composition. PRS were calculated using PRSice-2 [29] and GWAS summary statistics from blood cell count traits including monocytes, basophils, erythrocytes, platelets, lymphocytes, and neutrophils [30]. Variants with AD association test P-values < 5.0×10^-5^ and MAF > 0.01 from each GWAS for individuals with EA, African and Hispanic/Latino ancestry were selected for PRS calculation and then LD-clumped (r^2^ < 0.1). The adjusted TL values were then z-scaled within each ancestry group. The distribution of scaled TL across the total sample and within each ancestry group is shown in **Additional File 1: Fig. S3**. TL was dichotomized into long (TL > 0) and short (TL ≤ 0) groups within each ancestry group based on the median for the group. Characteristics of individuals in the long and short TL groups are shown in **Fig. 1** and **Additional File 1: Fig. S4.**

**Fig. 1.**
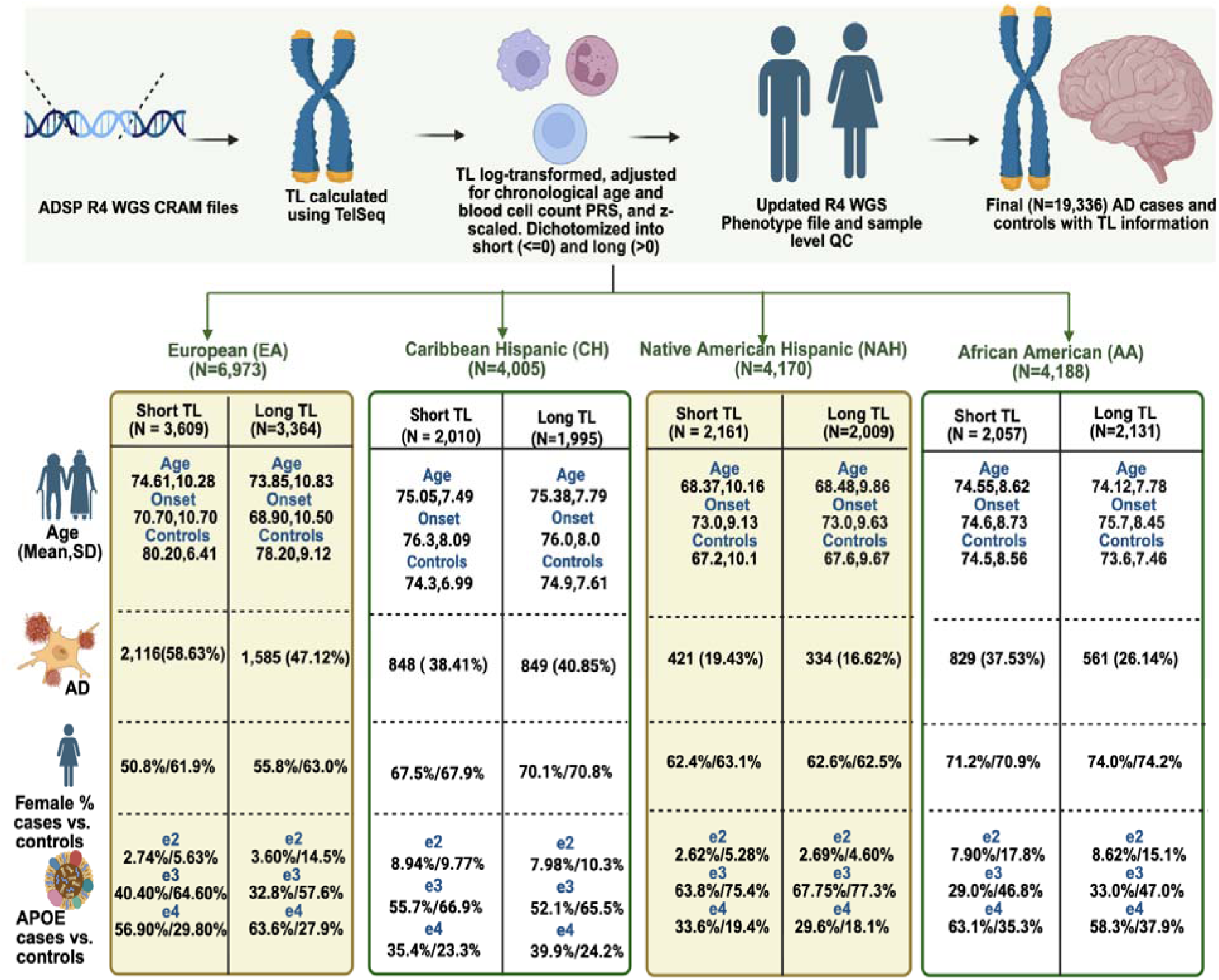
Study design and characteristics of the study participants across different ancestries. AD = Alzheimer disease; TL = telomere length. Created in BioRender. Khurshid, Z. (2026) https://BioRender.com/b2hkvsq

### Association of Telomere Length with AD

A linear regression model implemented in R (lm function) [31] was used to test the association of TL as the outcome with AD and covariates for sex, dosage of the *APOE* ε2 and ε4 alleles, and PRS for the six blood cell type proxies. Analyses were performed in the total sample and within each ancestry group. Genome-wide analyses testing the association of AD with the interaction between dichotomized TL and SNPs having MAC > 20 were performed in each ancestry group (EA = 7,554,931, CH = 12,052,554, NAH = 6,276,798, and AA = 13,782,316) using logistic regression models implemented in GENESIS including main effects for SNP and TL, an interaction term (SNP × TL), and covariates for age, sex, sequencing center, *APOE* ε2 and ε4 allele dosages, GRM, and the first 20 ancestry-specific PCs. We set thresholds at P < 5.0×10^-8^ for genome-wide significant and P < 1.0×10^-5^ for suggestive associations. Genomic control correction was applied as needed. Results across ancestry groups were combined using the METASOFT RE2 model [32]. Variants were annotated using SnpEFF (V4.3t) and Ensembl Variant Effect Predictor (GRCh38) [33,34] For each lead SNP, regional association plots were generated using the locuszoomr R package [35]. LD (r^2^) was computed from study genotypes as the squared Pearson correlation of alternate-allele dosages between each variant and the lead SNP within a 500 kb window.

### Differential Expression Analysis

Bulk RNA-Seq from the Religious Orders Study and Memory and Aging Project ROSMAP [36,37] were analyzed for participants who also had WGS data available (n = 483) across three brain regions: dorsolateral prefrontal cortex (DLPFC; 194 AD cases, 224 controls; 217 Long TL, 201 Short TL), posterior cingulate cortex (PCC; 101 AD cases, 152 controls; 113 Long TL, 140 Short TL), and anterior cingulate cortex (AC; 145 AD cases, 190 controls; 165 Long TL, 171 Short TL). Differential gene expression (DGE) was performed for the top results from the SNP x TL interaction GWAS analysis using DESeq2 [38]. The bulk RNA-seq data analysis model was adjusted for AD status, age of death, RNA integrity number (RIN), library batch, and sex with expression as outcome.

## Results

In the total sample, analysis of the model with all variables revealed that shorter TL was significantly associated with AD status (β = -0.048, *P* < 2×10^-16^), females had significantly longer TL than males (β = 0.009, *P* = 7.47×10^-5^), and dosage of *APOE* ε2 (β = 0.012, 3.40×10^-4^) and *APOE* ε4 (β = -0.005, *P* = 7.05×10^-3^) were associated with longer and shorter TL, respectively, compared to the ε3/ε3 genotype (**Table 2**). The strength of the association of AD and shorter TL was comparable in EA (β = -0.054, *P* < 2×10^-16^) and AA (β = -0.047, *P* < 2×10^-16^) individuals, but attenuated in NAH individuals (β = -0.012, *P* = 0.038). AD was not associated with TL in the CH group. Longer TL in females was evident in the EA (β = 0.009, *P* = 0.016), CH (β = 0.010, *P* = 0.032), and AA (β = 0.010, *P* = 0.050) groups. The association of longer TL with dosage of *APOE* ε4 was significant in the EA (β = 0.006, *P* = 0.044) and CH (β = 0.014, *P* = 4.5×10^-4^) groups, but the association with dosage of *APOE* ε2 was significant in the EA group only (β = 0.056, *P* < 2×10^-16^).

**Table 2.** Association of TL with AD, age, sex, *APOE* genotype, and cell type by ancestry.

| Total (n=19336) |  |  |  |
| --- | --- | --- | --- |
| | Effect ($\beta$) | Std.Error | P |
| Intercept | 0.966 | 0.137 | 1.91E-12 |
| Age | -0.005 | 0.000 | <2.00E-16 |
| AD | -0.048 | 0.002 | <2.00E-16 |
| Sex | 0.009 | 0.002 | 7.47E-05 |
| *APOE* ε2 | 0.012 | 0.003 | 3.40E-04 |
| *APOE* ε4 | -0.005 | 0.002 | 7.05E-03 |
| PRS BASO | -2.080 | 0.294 | 1.47E-12 |
| PRS EOS | -1.816 | 0.288 | 3.14E-10 |
| PRS MONO | 0.484 | 0.414 | 2.43E-01 |
| PRS LYM | -1.103 | 0.383 | 4.00E-03 |
| PRS NEU | -1.742 | 0.420 | 3.37E-05 |
| PRS PLT | -0.859 | 0.533 | 1.07E-01 |
| EA (n=6973) |  |  |  |
| | Effect ($\beta$) | Std.Error | P |
| Intercept | 1.475 | 0.236 | 4.45E-10 |
| Age | -0.006 | 0.000 | <2.00E-16 |
| AD | -0.054 | 0.004 | <2.00E-16 |
| Sex | 0.009 | 0.004 | 1.56E-02 |
| *APOE* ε2 | 0.056 | 0.006 | <2.00E-16 |
| *APOE* ε4 | 0.006 | 0.003 | 4.40E-02 |
| PRS BASO | -0.243 | 0.461 | 6.00E-01 |
| PRS EOS | 0.316 | 0.583 | 5.89E-01 |
| PRS MONO | 0.344 | 1.008 | 7.33E-01 |
| PRS LYM | -0.212 | 0.921 | 8.18E-01 |
| PRS NEU | 0.282 | 1.590 | 8.59E-01 |
| PRS PLT | -2.004 | 1.384 | 1.48E-01 |
| CH (n=4005) |  |  |  |
| | Effect ($\beta$) | Std.Error | P |
| Intercept | 1.458 | 0.244 | 2.49E-09 |
| Age | -0.003 | 0.000 | <2.00E-16 |
| AD | 0.002 | 0.005 | 7.23E-01 |
| Sex | 0.010 | 0.005 | 3.20E-02 |
| *APOE* ε2 | 0.012 | 0.007 | 7.30E-02 |
| *APOE* ε4 | 0.014 | 0.004 | 4.50E-04 |
| PRS BASO | -0.326 | 0.521 | 5.32E-01 |
| PRS EOS | -0.295 | 0.548 | 5.91E-01 |
| PRS MONO | 0.755 | 0.763 | 3.23E-01 |
| PRS LYM | -1.667 | 0.700 | 1.70E-02 |
| PRS NEU | -0.266 | 0.785 | 7.35E-01 |
| PRS PLT | -1.373 | 0.958 | 1.52E-01 |
| NAH (n=4170) |  |  |  |
| | Effect ($\beta$) | Std.Error | P |
| Intercept | 0.381 | 0.341 | 2.65E-01 |
| Age | -0.002 | 0.000 | <2.00E-16 |
| AD | -0.012 | 0.005 | 3.80E-02 |
| Sex | 0.002 | 0.005 | 5.95E-01 |
| *APOE* ε2 | -0.011 | 0.010 | 2.60E-01 |
| *APOE* ε4 | -0.008 | 0.005 | 1.25E-01 |
| PRS BASO | -2.207 | 0.692 | 1.40E-03 |
| PRS EOS | -2.842 | 0.740 | 1.20E-04 |
| PRS MONO | -0.822 | 1.312 | 5.31E-01 |
| PRS LYM | 2.409 | 1.283 | 6.00E-02 |
| PRS NEU | 2.118 | 1.726 | 2.19E-01 |
| PRS PLT | -0.020 | 1.619 | 9.92E-01 |
| AA (n=4188) |  |  |  |
| | Effect ($\beta$) | Std.Error | P |
| Intercept | 2.010 | 0.314 | 1.64E-10 |
| Age | -0.006 | 0.000 | <2.00E-16 |
| AD | -0.047 | 0.005 | <2.00E-16 |
| Sex | 0.010 | 0.005 | 5.00E-02 |
| *APOE* ε2 | -0.006 | 0.006 | 2.56E-01 |
| *APOE* ε4 | 0.000 | 0.004 | 9.16E-01 |
| PRS BASO | 0.436 | 0.705 | 5.36E-01 |
| PRS EOS | 0.109 | 0.457 | 8.11E-01 |
| PRS MONO | 0.569 | 0.594 | 3.39E-01 |
| PRS LYM | -0.726 | 0.529 | 1.70E-01 |
| PRS NEU | -1.472 | 0.558 | 8.00E-03 |
| PRS PLT | 0.832 | 0.730 | 2.54E-01 |

AD = Alzheimer disease; TL = telomere length; EA = European; CH = Caribbean Hispanic; NAH = Native American Hispanic; AA = African American; BASO = basophils; EOS = erythrocytes; MONO = monocytes; LYM = lymphocytes; NEU = neutrophils; PLT = platelets

An AD GWAS for the interaction between dichotomized TL and genetic variants in the EA group revealed genome-wide significant (GWS) interactions involving SNPs spanning several loci on chromosome 5 including *SEMA6A (*rs976030371*, P*=1.42×10^-8^; rs950795839, *P*=1.45×10^-8^), *RP11-249M12.2* (rs1005099552, *P*=1.45×10^-8^), *CTC-472C24.1* (rs934213781, *P*=1.45×10^-8^), and *LOC105378654 (*rs537146885*, P*=4.17×10^-8^), as well as in regions between *CTC-472C24.1* and *RP11-492L8.1* (5:116851056, *P*=1.45×10^-8^), *IL15* and *INPP4B* (rs549471753*, P*=1.77×10^-8^), in *LOC105378654 (*rs537146885*, P*=4.17×10^-8^), and between *LINC02208* and *LINC02216* (rs1277036803, *P*=4.60×10^-8^) (**Table 3, Additional File 2: Table S2, Additional File 1: Fig. S5**). In the NAH group, GWS TL interactions were observed with *BSN* intronic SNP rs891582930 (*P*=3.26×10^-8^) and a missense variant in *MST1* (rs138155786, *P*=3.26×10^-8^) located 11 kb from *BSN* (**Table 3, Additional File 2: Table S2, Additional File 1: Fig. S7**). No TL x SNP interactions were GWS in the CH and AA groups, although several reached the suggestive significance level including *MIR646HG* intronic SNP rs141837617 (*P*=9.87×10^-8^) in the CH group and *ASIC2* intronic SNP rs34446836 (*P*=3.02×10^-7^) in the AA group (**Additional File 2: Table S2, Additional File 1: Fig. S7 and Fig. S8**). Locus zoom plots showed that the five GWS SNPs in the *SEMA6A* region are high in high LD, noting that the SEMA6A variant is exonic and the four others are non-coding. Similarly, the *BSN* and *MST1* SNPs appear to be in complete LD (**Additional File 1: Fig. S9**). Meta-analysis of results across population groups revealed that the interaction between TL and rs537146885 was more significant *(P*=3.36×10^-9^) due solely to the added contribution from the CH group (**Table 3, Additional File 2: Table S3**). GWS interactions were also observed between TL and three SNPs located between *CTD-2160D9.*1 and EEF1A1P20 including rs146719444 *(P*=9.55×10^-9^), rs138582861 (*P*=1.19×10^-8^), and rs147421008 (*P*=3.58×10^-8^) with evidence in AA and CHs. These variants were very rare or absent in the EA and NAH group. The interaction between TL and *TBC1D22A* intronic SNP rs117006149 was GWS (*P*=1.06×10^-8^) with most support in the EA group but also significant in both Hispanic groups. We also found a GWS interaction with *PLK1* rs955713021 (*P*=3.28×10^-8^) primarily due to evidence in the EA group.

**Table 3.** Genome-wide significant SNP × TL interactions by ancestry.

| EA: short TL=3609, long TL=3364 |  |  |  |  |  |  |  |  |  |
| --- | --- | --- | --- | --- | --- | --- | --- | --- | --- |
| Variant ID | Position | Gene | Function | P | MAF Cases | MAF Control | MAF Short TL | MAF Long TL |  |
| rs976030371 | 5:116540412:A:G | *SEMA6A* | Intron | 1.42E-08 | 0.0007 | 0.0035 | 0.0024 | 0.0016 |  |
| rs950795839 | 5:116539305:G:A | *SEMA6A* | Intron | 1.45E-08 | 0.0007 | 0.0035 | 0.0024 | 0.0016 |  |
| rs1005099552 | 5:116759521:A:C | *RP11-249M12.2* | Intron | 1.45E-08 | 0.0007 | 0.0035 | 0.0024 | 0.0016 |  |
| rs934213781 | 5:116823313:T:C | *CTC-472C24.1* | Intron | 1.45E-08 | 0.0007 | 0.0035 | 0.0024 | 0.0016 |  |
|  | 5:116851056:G:A | *CTC-472C24.1-RP11-492L8.1* | Intergenic region | 1.47E-08 | 0.0007 | 0.0035 | 0.0024 | 0.0016 |  |
| rs549471753 | 4:141903346:C:T | *IL15-INPP4B* | Intergenic region | 1.77E-08 | 0.0016 | 0.0040 | 0.0033 | 0.0021 |  |
| rs537146885 | 1:38198857:C:T | *LOC105378654* | Intron | 4.17E-08 | 0.0018 | 0.0038 | 0.0026 | 0.0028 |  |
| rs1277036803 | 5:118570960:G:C | *LINC02208-LINC02216* | Upstream gene | 4.60E-08 | 0.0007 | 0.0029 | 0.0019 | 0.0015 |  |
| NAH: short TL=2161, long TL=2009 |  |  |  |  |  |  |  |  |  |
| rs891582930 | 3:49614753:A:T | *BSN* | Intron | 3.26E-08 | 0.0027 | 0.0025 | 0.0005 | 0.0047 |  |
| rs138155786 | 3:49684378:C:T | *MST1* | Missense | 3.26E-08 | 0.0027 | 0.0025 | 0.0005 | 0.0047 |  |
| Total (EA + NAH + CH + AA): short TL=9837, long TL=9499 |  |  |  |  |  |  |  |  |  |
| Variant ID | Position | Gene | Function | P | MAF Cases | MAF Control | MAF Short TL | MAF Long TL | Effect Direction* |
| rs537146885 | 1:38198857:C:T | *LOC105378654* | Intron | 3.36E-09 | 0.0016 | 0.0022 | 0.0015 | 0.0025 | --?? |
| rs146719444 | 5:99959505:G:A | *CTD-2160D9.1-EEF1A1P20* | Intergenic region | 9.55E-09 | 0.0070 | 0.0076 | 0.0076 | 0.0072 | ?-?- |
| rs138582861 | 5:99982763:CAGTA:C | *CTD-2160D9.1-EEF1A1P20* | Intergenic region | 1.19E-08 | 0.0069 | 0.0076 | 0.0075 | 0.0072 | ?-?- |
| rs147421008 | 5:99974699:C:T | *CTD-2160D9.1-EEF1A1P20* | Intergenic region | 3.58E-08 | 0.0069 | 0.0074 | 0.0074 | 0.0070 | ?-?- |
| rs117006149 | 22:46854696:C:T | *TBC1D22A* | Intron | 1.06E-08 | 0.0145 | 0.0105 | 0.0117 | 0.0124 | +++? |
| rs955713021 | 16:23682991:T:C | *PLK1* | Intron | 3.28E-08 | 0.0048 | 0.0059 | 0.0043 | 0.0067 | --+? |

EA = European; NAH = Native American Hispanic; CH = Caribbean Hispanic; AA = African American; MAF = minor allele frequency; TL = telomere length;
\* Direction order of ancestries: EA, CH, NAH, AA

DGE analysis revealed significantly lower expression in the PCC of *SEMA6A* (log₂FC = - 0.16, *P_adj_*=0.033) and *MIR646HG* (log₂FC = -0.36, *P_adj_*=0.038) and higher expression of *PLK1* (log₂FC = 0.32, *P_adj_*=0.033) in AD cases compared to controls with long TL (**Fig. 2, Additional File 2: Table S4**). Among individuals with short TL, expression in the DLPFC was significantly lower for *BSN* (log₂FC = −0.16, *P_adj_*=0.0069) and *ASIC2* (log₂FC = -0.09, *P_adj_*=0.030) and higher for *MST1* (log₂FC = 0.13, *P_adj_*=0.013) in AD cases compared to controls. Notably, each of these genes was not significantly differentially expressed in the corresponding short or long TL group, and the pattern of expression was often in the opposite direction.

**Fig. 2.**
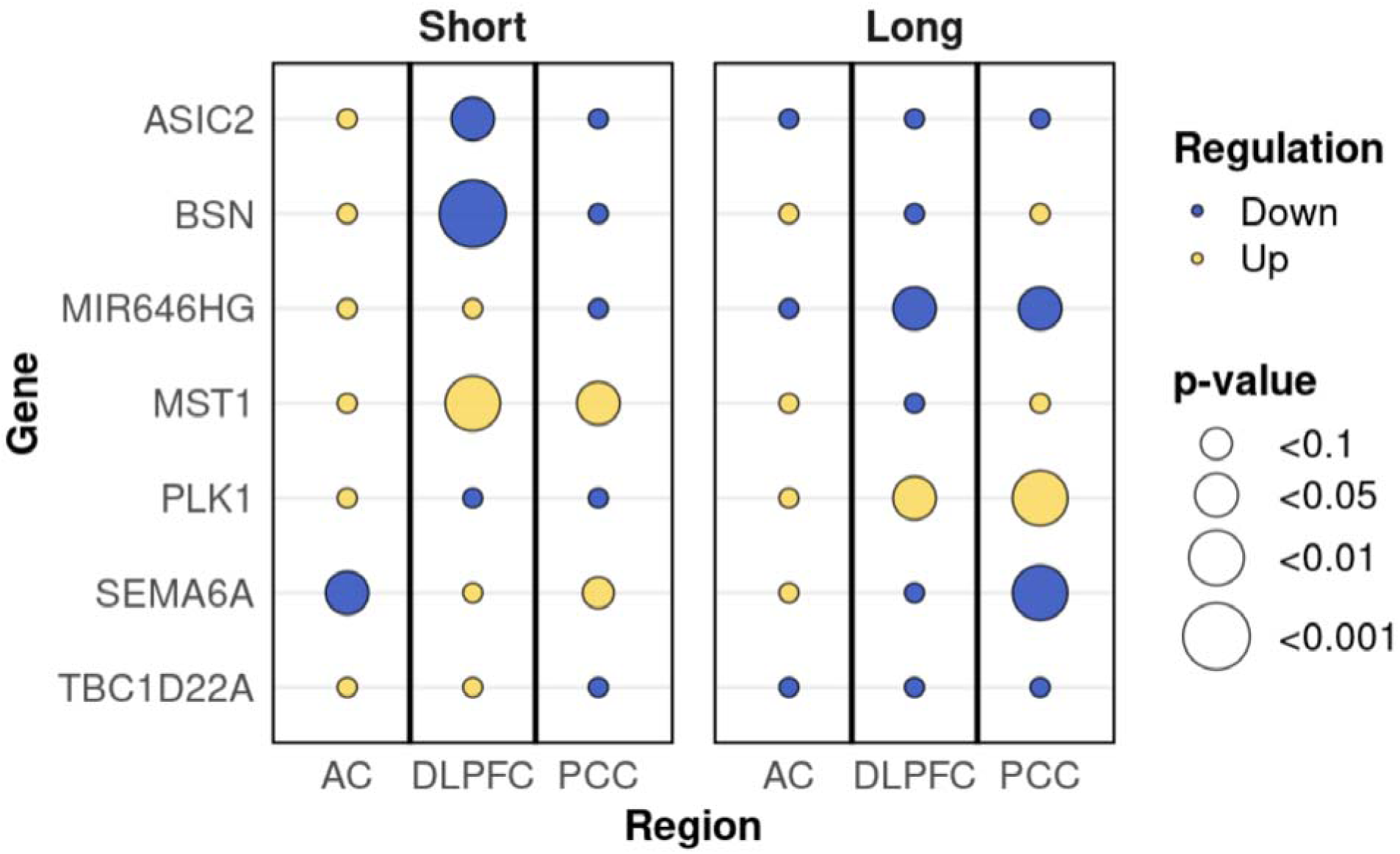
Differential expression between AD cases and controls for genes containing variants in significant SNP×TL interactions. AD = Alzheimer disease; TL = telomere length; AC = anterior cingulate cortex; DLPFC = dorsolateral prefrontal cortex; PCC = posterior cingulate cortex.

## Discussion

In this study of TL measured in WGS data obtained from a multi-ancestry sample of 19,336 ADSP participants, we confirmed previous findings showing a correlation between shorter TL and increased risk of AD and observed that dosages of the *APOE* ε2 and ε4 alleles compared to the ε3/ε3 genotype were significantly associated with longer TL [17,39,40]. We also observed known sex-based TL differences with females having longer TL on average than their male counterparts [41]. Our genome-wide scan for genetic variants modifying the association of TL with AD risk identified in EA individuals GWS associations with rare variants in a chromosome 5 locus including *SEMA6A, LOC105378654*, and intergenic regions between *IL15* and *INPP4B* and between *LINC02208* and *LINC02215*. In the NAH group, GWS interactions were observed with an intronic variant in *BSN* and a missense variant in a neighboring gene, *MST1*. No GWS TL × SNP interactions were detected in the CH and AA groups, although several reached the suggestive significance level. Meta-analysis of GWAS results across four ancestry groups revealed additional GWS interactions involving variants in *LOC105378654, TBC1D22A* and *PLK1.* All seven protein coding genes in this group were differentially expressed between AD cases and controls in the PCC or DLPFC among individuals with short or long TL, but not both, suggesting that the mechanism underlying the significant TL × SNP interactions involves effects of TL on protein abundance in brain regions most affected by AD.

The protein encoded by *SEMA6A* (Semaphorin 6A) is involved in cell signaling, axon guidance and cell migration and plays a role in nervous system development [42,43]. Semaphorins are increasingly recognized for their roles in neuroinflammation, synaptic remodeling, and blood–brain barrier dynamics which are relevant to neurodegeneration. Multiple semaphorins have been mechanistically linked to AD-related processes including regulating the levels of Aβ, hyperphosphorylation of tau protein, and Reelin [44,45] and adult hippocampal connectivity and memory processes [46]. *SEMA6A* and Plexin A2/A4 signaling determines settling position of superficial layer neurons and Plexin A4 mediates amyloid-β (Aβ)-induced tau pathology in AD [47,48,49]. Expression of *SEMA6A* was reduced in the PCC of AD cases with long TL suggesting TL-dependent effects on neurodevelopment and inflammatory signaling.

*BSN* (Bassoon Presynaptic Cytomatrix Protein) has roles in the structural organization of the presynaptic cytoskeleton, maintaining synaptic integrity and regulating neurotransmitter release [50]. Variants in *BSN* have been linked to multiple neuropsychiatric and neurodegenerative disorders, including schizophrenia, PSP, multiple system atrophy (MSA), epilepsy, bipolar disorder, and Parkinson’s disease [51]. One study showed that downregulation of *BSN* reduces tau seeding, spreading, and overall neurotoxicity in AD mouse models [52]. The *BSN* intronic variant involved in the GWS interaction with TL might be functionally relevant because it has a Combined Annotation Dependent Depletion (CADD) PHRED-scaled score (GRCh38, v1.7) [53] of 11.84, which is within the top 10% of deleterious variants, but this needs to be confirmed experimentally. In our analysis, it showed a significant SNP × TL interaction with AD, with a risk effect in the shorter TL group. In the DLPFC, it was downregulated in AD cases with short TL indicating that telomere-associated synaptic dysfunction may contribute to tau-related pathology and synaptic loss in AD. However, the biological signal underlying the association at this locus is more strongly tied to the highly correlated variant in the adjacent gene, *MST1*. *MST1* (Macrophage-stimulating 1) encodes macrophage-stimulating protein (MSP), a ligand for the RON (Recepteur d’Origine Nantais) receptor that modulates anti-inflammatory signaling. Impaired MSP–RON signaling in microglia has been shown to enhance neuroinflammation in the central nervous system, a key process in AD pathogenesis [54,55,56]. MSP is also linked to metabolic syndrome which is a known AD risk factor [57,58]. The PHRED score for the *MST1* missense variant is 24.2, indicating that it is very likely deleterious and within the top 1% of potentially pathogenic variants genome-wide. We observed a significant association between AD and the *MST1* variant among individuals with shorter TL. *MST1* was upregulated in the DLPC of AD cases with short TL suggesting that inflammatory activation of the MSP–RON pathway may exacerbate neurodegenerative processes under telomere stress.

An abnormally high level of *PLK1* (Polo like Kinase 1) has been observed in brains of pathologically diagnosed AD cases and its inhibition reduced Aβ -induced neuronal cell death [59,60]. We identified a *PLK1* variant that conferred increased risk of AD among individuals with long TL and *PLK1* expression was higher in the PCC of AD cases with long TL consistent with excessive *PLK1* activity contributing to neurodegenerative vulnerability. The relevance to AD of other genes implicated in this study is less clear.

Mechanisms underlying the observed significant TL × SNP interactions are not immediately apparent; however, genetic variants and TL may jointly influence AD risk through pathways sensitive to cellular stress, inflammation, and neuronal homeostasis. Shorter TL is associated with higher replicative and oxidative stress and thus may activate immune and stress-response pathways leading to inflammation, which is a hallmark of AD [61,62,63,64]. Several findings from our study support this model. *SEMA6A*, induced under inflammatory conditions, may help preserve microglial and synaptic integrity, whereas increased expression of *MST1* under sustained inflammatory pressure may promote neurodegeneration. Short TL in combination with *BSN* downregulation may increase synaptic vulnerability and impaired neurotransmission. In contrast, longer TL which typically reflects reduced cellular stress or delayed senescence may predispose to dysregulation of proliferative and synaptic maintenance processes. For example, *PLK1* upregulation among individuals with long TL may promote aberrant neuronal cell cycle reactivation. Notably, both *APOE* ε2 and ε4 alleles were associated with longer TL in our cohort, but likely via distinct mechanisms. The ε2 association aligns with its neuroprotective and anti-inflammatory effects [65] and the significantly higher ε2 frequency among centenarians [66] potentially reflects preserved telomere maintenance. Consistent with this idea, the ε2 protective effect was observed in both short and high TL groups but was stronger among individuals with longer TL (β = -0.32, *P*=1×10^-5^) than shorter TL (β = -0.21, *P*= 4×10^-3^), suggesting that telomere preservation may enhance ε2-mediated resilience against AD. The association of long TL with ε4 is counterintuitive because of the strong effect of ε4 on AD risk. However, prior studies indicate that ε4 carriers tend to have longer TL and longer telomeres among ε4 carriers predicts worse episodic memory [40] and faster cognitive decline particularly among ε4 homozygotes [67].

Ancestry-specific differences in TL × SNP interactions likely reflect variation in allele frequencies, established genetic differences, and environmental factors that influence TL across populations [18,68]. Differences in sample size and power, particularly for rare variants in CH and AA groups, also contribute to why GWS signals were detected in EA and NAH but only suggestive associations appeared in other ancestries.

Our study has several notable strengths. Investigation of the interaction between genetic variants and TL offered an opportunity to increase discovery of rare variants whose effect on AD risk is modulated by TL and explore factors that might elucidate mechanisms linking TL to AD. A key strength of our study is its multi-ancestry design, which addresses a critical gap in TL studies in AD that have historically focused primarily on EA populations. We also accounted for confounders including *APOE* genotype, blood cell counts, age, sex, and population structure, reducing the likelihood that observed associations were due to uncontrolled bias. Many of our most significant association findings were supported by analyses showing differential gene expression between AD cases and controls. Furthermore, our study presents a scalable framework for evaluating the role of TL as a biological modifier in complex diseases, offering a resource for future studies exploring gene × environment or gene × aging interactions.

We also acknowledge several study limitations. Since we used computationally inferred TL rather than direct laboratory measurements, there may have been residual measurement error or unaccounted technical variation. TL was estimated from whole-genome sequencing data using TelSeq, which indicates TL at a single time point corresponding to the blood draw and does not capture longitudinal changes. Therefore, we were unable to assess dynamic telomere attrition over time, which may offer further insights into biological aging and disease progression. In addition, actual blood cell subtype counts were not available, and we relied on PRS to adjust for potential confounding from blood composition, which may not fully capture cell-type heterogeneity. Although our overall sample size was substantial, it was underpowered for detecting modest effect associations, particularly among rare variants or in the smaller non-EA ancestry groups. Most of our top signals involved rare variants, which are inherently more difficult to replicate or validate across datasets and, thus, these findings should be considered exploratory and interpreted with caution.

## Conclusions

In summary, we identified several novel AD-associated variants whose effects are modulated by TL, providing valuable insights into the relationship between telomere biology and AD pathogenesis. These results expand the genetic landscape of TL-associated risk and provide a foundation for future studies aimed at identifying new therapeutic targets that integrate telomere biology with neurodegeneration.

## Data Availability

Genetic and phenotype data for the ADSP dataset used in this study are available through the National Institute on Aging Alzheimer's Disease Data Storage Site (NIAGADS) (https://dss.niagads.org/datasets/ng00067/)

## List of abbreviations

AA: African American
Aβ: Amyloid-β
AC: Anterior cingulate cortex
AD: Alzheimer disease
ADSP: Alzheimer’s Disease Sequencing Project
BSN: Bassoon Presynaptic Cytomatrix Protein
CADD: Combined Annotation Dependent Depletion
CBD: Corticobasal Degeneration
CDR: Clinical Dementia Rating
CH: Caribbean Hispanic
DGE: Differential gene expression
DLPFC: Dorsolateral prefrontal cortex
EA: European
GATK: Genome Analysis Toolkit
GCAD: Genome Center for Alzheimer’s Disease
GENESIS: GENetic EStimation and Inference in Structured samples
GMM: Gaussian Mixture Model
GRM: Genetic relationship matrix
GWS: Genome-wide significant
GWAS: Genome-wide association studies
gVCF: Genomic VCF
LD: Linkage disequilibrium
MAC: Minor allele count
MAF: Minor allele frequency
MSA: Multiple system atrophy
MST1: Macrophage-stimulating 1
MSP: Macrophage-stimulating protein
NAH: Native American Hispanic
NIAGADS: National Institute on Aging Genetics of Alzheimer Disease Data Storage site
PC: Principal component
PCC: Posterior cingulate cortex
pVCF: Project-level VCF
PLK1: Polo like Kinase 1
PRS: Polygenic risk score
PSP: Progressive Supranuclear Palsy
QC: Quality control
R4: Release 4 (dataset)
RIN: RNA integrity number
RON: Recepteur d’Origine Nantais
SA: South Asian
SEMA6A: Semaphorin 6A
TERT: Telomerase reverse transcriptase
TL: Telomere length
VCPA: Variant Calling Pipeline and data management tool
WGS: Whole genome sequencing

## Declarations

### Ethics approval and consent to participate

Not applicable.

### Consent for publication

Not applicable

### Authors’ contributions

Z.K. contributed to conceptualization, methodology, formal analysis, data curation, visualization, and writing of the original draft. J.J.F. contributed to data curation, software, and writing–review and editing. T.T., O.O., and C.Z. contributed to data curation. E.M., W.B., M.P.-V., G.S., and J.H. contributed to funding acquisition and writing–review and editing. L.-S.W. provided resources. K.L. contributed to data analysis supervision and writing–review and editing. X.Z. contributed to conceptualization, methodology, supervision, and writing–review and editing. L.F. contributed to conceptualization, methodology, supervision, project administration, writing–review and editing, and funding acquisition. All authors read and approved the final manuscript.

## Acknowledgments

This study was supported by National Institute on Aging grants R01-AG048927, U01-AG058654, U01-AG062602, P30-AG072978, U19-AG068753, U01-AG072577, R01-AG080810, and U01-AG082665.

The Alzheimer’s Disease Sequencing Project (ADSP) is comprised of two Alzheimer’s Disease (AD) genetics consortia and three National Human Genome Research Institute (NHGRI) funded Large Scale Sequencing and Analysis Centers (LSAC). The two AD genetics consortia are the Alzheimer’s Disease Genetics Consortium (ADGC) funded by NIA (U01 AG032984), and the Cohorts for Heart and Aging Research in Genomic Epidemiology (CHARGE) funded by NIA (R01 AG033193), the National Heart, Lung, and Blood Institute (NHLBI), other National Institute of Health (NIH) institutes and other foreign governmental and non-governmental organizations. The Discovery Phase analysis of sequence data is supported through UF1AG047133 (to Drs. Schellenberg, Farrer, Pericak-Vance, Mayeux, and Haines); U01AG049505 to Dr. Seshadri; U01AG049506 to Dr. Boerwinkle; U01AG049507 to Dr. Wijsman; and U01AG049508 to Dr. Goate and the Discovery Extension Phase analysis is supported through U01AG052411 to Dr. Goate, U01AG052410 to Dr. Pericak-Vance and U01 AG052409 to Drs. Seshadri and Fornage.

Sequencing for the Follow Up Study (FUS) is supported through U01AG057659 (to Drs. PericakVance, Mayeux, and Vardarajan) and U01AG062943 (to Drs. Pericak-Vance and Mayeux). Data generation and harmonization in the Follow-up Phase is supported by U54AG052427 (to Drs. Schellenberg and Wang). The FUS Phase analysis of sequence data is supported through U01AG058589 (to Drs. Destefano, Boerwinkle, De Jager, Fornage, Seshadri, and Wijsman), U01AG058654 (to Drs. Haines, Bush, Farrer, Martin, and Pericak-Vance), U01AG058635 (to Dr. Goate), RF1AG058066 (to Drs. Haines, Pericak-Vance, and Scott), RF1AG057519 (to Drs. Farrer and Jun), R01AG048927 (to Dr. Farrer), and RF1AG054074 (to Drs. Pericak-Vance and Beecham).

The ADGC cohorts include: Adult Changes in Thought (ACT) (U01 AG006781, U19 AG066567), the Alzheimer’s Disease Research Centers (ADRC) (P30 AG062429, P30 AG066468, P30 AG062421, P30 AG066509, P30 AG066514, P30 AG066530, P30 AG066507, P30 AG066444, P30 AG066518, P30 AG066512, P30 AG066462, P30 AG072979, P30 AG072972, P30 AG072976, P30 AG072975, P30 AG072978, P30 AG072977, P30 AG066519, P30 AG062677, P30 AG079280, P30 AG062422, P30 AG066511, P30 AG072946, P30 AG062715, P30 AG072973, P30 AG066506, P30 AG066508, P30 AG066515, P30 AG072947, P30 AG072931, P30 AG066546, P20 AG068024, P20 AG068053, P20 AG068077, P20 AG068082, P30 AG072958, P30 AG072959), the Chicago Health and Aging Project (CHAP) (R01 AG11101, RC4 AG039085, K23 AG030944), Indiana Memory and Aging Study (IMAS) (R01 AG019771), Indianapolis Ibadan (R01 AG009956, P30 AG010133), the Memory and Aging Project (MAP) ( R01 AG17917), Mayo Clinic (MAYO) (R01 AG032990, U01 AG046139, R01 NS080820, RF1 AG051504, P50 AG016574), Mayo Parkinson’s Disease controls (NS039764, NS071674, 5RC2HG005605), University of Miami (R01 AG027944, R01 AG028786, R01 AG019085, IIRG09133827, A2011048), the Multi-Institutional Research in Alzheimer’s Genetic Epidemiology Study (MIRAGE) (R01 AG09029, R01 AG025259), the National Centralized Repository for Alzheimer’s Disease and Related Dementias (NCRAD) (U24 AG021886), the National Institute on Aging Late Onset Alzheimer’s Disease Family Study (NIA-LOAD) (U24 AG056270), the Religious Orders Study (ROS) (P30 AG10161, R01 AG15819), the Texas Alzheimer’s Research and Care Consortium (TARCC) (funded by the Darrell K Royal Texas Alzheimer’s Initiative), Vanderbilt University/Case Western Reserve University (VAN/CWRU) (R01 AG019757, R01 AG021547, R01 AG027944, R01 AG028786, P01 NS026630, and Alzheimer’s Association), the Washington Heights-Inwood Columbia Aging Project (WHICAP) (RF1 AG054023), the University of Washington Families (VA Research Merit Grant, NIA: P50AG005136, R01AG041797, NINDS: R01NS069719), the Columbia University Hispanic Estudio Familiar de Influencia Genetica de Alzheimer (EFIGA) (RF1 AG015473), the University of Toronto (UT) (funded by Wellcome Trust, Medical Research Council, Canadian Institutes of Health Research), and Genetic Differences (GD) (R01 AG007584).

The CHARGE cohorts are supported in part by National Heart, Lung, and Blood Institute (NHLBI) infrastructure grant HL105756 (Psaty), RC2HL102419 (Boerwinkle) and the neurology working group is supported by the National Institute on Aging (NIA) R01 grant AG033193. The CHARGE cohorts participating in the ADSP include the following: Austrian Stroke Prevention Study (ASPS), ASPS-Family study, and the Prospective Dementia Registry-Austria (ASPS/PRODEM-Aus), the Atherosclerosis Risk in Communities (ARIC) Study, the Cardiovascular Health Study (CHS), the Erasmus Rucphen Family Study (ERF), the Framingham Heart Study (FHS), and the Rotterdam Study (RS). ASPS is funded by the Austrian Science Fond (FWF) grant number P20545-P05 and P13180 and the Medical University of Graz. The ASPS-Fam is funded by the Austrian Science Fund (FWF) project I904), the EU Joint Programme – Neurodegenerative Disease Research (JPND) in frame of the BRIDGET project (Austria, Ministry of Science) and the Medical University of Graz and the Steiermärkische Krankenanstalten Gesellschaft. PRODEM-Austria is supported by the Austrian Research Promotion agency (FFG) (Project No. 827462) and by the Austrian National Bank (Anniversary Fund, project 15435. ARIC research is carried out as a collaborative study supported by NHLBI contracts (HHSN268201100005C, HHSN268201100006C, HHSN268201100007C, HHSN268201100008C, HHSN268201100009C, HHSN268201100010C, HHSN268201100011C, and HHSN268201100012C). Neurocognitive data in ARIC is collected by U01 2U01HL096812, 2U01HL096814, 2U01HL096899, 2U01HL096902, 2U01HL096917 from the NIH (NHLBI, NINDS, NIA and NIDCD), and with previous brain MRI examinations funded by R01-HL70825 from the NHLBI. CHS research was supported by contracts HHSN268201200036C, HHSN268200800007C, N01HC55222, N01HC85079, N01HC85080, N01HC85081, N01HC85082, N01HC85083, N01HC85086, and grants U01HL080295 and U01HL130114 from the NHLBI with additional contribution from the National Institute of Neurological Disorders and Stroke (NINDS). Additional support was provided by R01AG023629, R01AG15928, and R01AG20098 from the NIA. FHS research is supported by NHLBI contracts N01-HC-25195 and HHSN268201500001I. This study was also supported by additional grants from the NIA (R01s AG054076, AG049607 and AG033040 and NINDS (R01 NS017950). The ERF study as a part of EUROSPAN (European Special Populations Research Network) was supported by European Commission FP6 STRP grant number 018947 (LSHG-CT-2006-01947) and also received funding from the European Community’s Seventh Framework Programme (FP7/2007-2013)/grant agreement HEALTH-F4-2007-201413 by the European Commission under the programme “Quality of Life and Management of the Living Resources” of 5th Framework Programme (no. QLG2-CT-2002-01254). High-throughput analysis of the ERF data was supported by a joint grant from the Netherlands Organization for Scientific Research and the Russian Foundation for Basic Research (NWO-RFBR 047.017.043). The Rotterdam Study is funded by Erasmus Medical Center and Erasmus University, Rotterdam, the Netherlands Organization for Health Research and Development (ZonMw), the Research Institute for Diseases in the Elderly (RIDE), the Ministry of Education, Culture and Science, the Ministry for Health, Welfare and Sports, the European Commission (DG XII), and the municipality of Rotterdam. Genetic data sets are also supported by the Netherlands Organization of Scientific Research NWO Investments (175.010.2005.011, 911-03-012), the Genetic Laboratory of the Department of Internal Medicine, Erasmus MC, the Research Institute for Diseases in the Elderly (014-93-015; RIDE2), and the Netherlands Genomics Initiative (NGI)/Netherlands Organization for Scientific Research (NWO) Netherlands Consortium for Healthy Aging (NCHA), project 050-060-810. All studies are grateful to their participants, faculty and staff. The content of these manuscripts is solely the responsibility of the authors and does not necessarily represent the official views of the National Institutes of Health or the U.S. Department of Health and Human Services.

The FUS cohorts include: the Alzheimer’s Disease Research Centers (ADRC) (P30 AG062429, P30 AG066468, P30 AG062421, P30 AG066509, P30 AG066514, P30 AG066530, P30 AG066507, P30 AG066444, P30 AG066518, P30 AG066512, P30 AG066462, P30 AG072979, P30 AG072972, P30 AG072976, P30 AG072975, P30 AG072978, P30 AG072977, P30 AG066519, P30 AG062677, P30 AG079280, P30 AG062422, P30 AG066511, P30 AG072946, P30 AG062715, P30 AG072973, P30 AG066506, P30 AG066508, P30 AG066515, P30 AG072947, P30 AG072931, P30 AG066546, P20 AG068024, P20 AG068053, P20 AG068077, P20 AG068082, P30 AG072958, P30 AG072959), Alzheimer’s Disease Neuroimaging Initiative (ADNI) (U19AG024904), Amish Protective Variant Study (RF1AG058066), Cache County Study (R01AG11380, R01AG031272, R01AG21136, RF1AG054052), Case Western Reserve University Brain Bank (CWRUBB) (P50AG008012), Case Western Reserve University Rapid Decline (CWRURD) (RF1AG058267, NU38CK000480), CubanAmerican Alzheimer’s Disease Initiative (CuAADI) (3U01AG052410), Estudio Familiar de Influencia Genetica en Alzheimer (EFIGA) (5R37AG015473, RF1AG015473, R56AG051876), Genetic and Environmental Risk Factors for Alzheimer Disease Among African Americans Study (GenerAAtions) (2R01AG09029, R01AG025259, 2R01AG048927), Gwangju Alzheimer and Related Dementias Study (GARD) (U01AG062602), Hillblom Aging Network (2014-A-004-NET, R01AG032289, R01AG048234), Hussman Institute for Human Genomics Brain Bank (HIHGBB) (R01AG027944, Alzheimer’s Association “Identification of Rare Variants in Alzheimer Disease”), Ibadan Study of Aging (IBADAN) (5R01AG009956), Longevity Genes Project (LGP) and LonGenity (R01AG042188, R01AG044829, R01AG046949, R01AG057909, R01AG061155, P30AG038072), Mexican Health and Aging Study (MHAS) (R01AG018016), Multi-Institutional Research in Alzheimer’s Genetic Epidemiology (MIRAGE) (2R01AG09029, R01AG025259, 2R01AG048927), Northern Manhattan Study (NOMAS) (R01NS29993), Peru Alzheimer’s Disease Initiative (PeADI) (RF1AG054074), Puerto Rican 1066 (PR1066) (Wellcome Trust (GR066133/GR080002), European Research Council (340755)), Puerto Rican Alzheimer Disease Initiative (PRADI) (RF1AG054074), Reasons for Geographic and Racial Differences in Stroke (REGARDS) (U01NS041588), Research in African American Alzheimer Disease Initiative (REAAADI) (U01AG052410), the Religious Orders Study (ROS) (P30 AG10161, P30 AG72975, R01 AG15819, R01 AG42210), the RUSH Memory and Aging Project (MAP) (R01 AG017917, R01 AG42210Stanford Extreme Phenotypes in AD (R01AG060747), University of Miami Brain Endowment Bank (MBB), University of Miami/Case Western/North Carolina A&T African American (UM/CASE/NCAT) (U01AG052410, R01AG028786), Wisconsin Registry for Alzheimer’s Prevention (WRAP) (R01AG027161 and R01AG054047), Mexico-Southern California Autosomal Dominant Alzheimer’s Disease Consortium (R01AG069013), Center for Cognitive Neuroscience and Aging (R01AG047649), and the A4 Study (R01AG063689, U19AG010483 and U24AG057437).

The four LSACs are: the Human Genome Sequencing Center at the Baylor College of Medicine (U54 HG003273), the Broad Institute Genome Center (U54HG003067), The American Genome Center at the Uniformed Services University of the Health Sciences (U01AG057659), and the Washington University Genome Institute (U54HG003079). Genotyping and sequencing for the ADSP FUS is also conducted at John P. Hussman Institute for Human Genomics (HIHG) Center for Genome Technology (CGT).

Biological samples and associated phenotypic data used in primary data analyses were stored at Study Investigators institutions, and at the National Centralized Repository for Alzheimer’s Disease and Related Dementias (NCRAD, U24AG021886) at Indiana University funded by NIA. Associated Phenotypic Data used in primary and secondary data analyses were provided by Study Investigators, the NIA funded Alzheimer’s Disease Centers (ADCs), and the National Alzheimer’s Coordinating Center (NACC, U24AG072122) and the National Institute on Aging Genetics of Alzheimer’s Disease Data Storage Site (NIAGADS, U24AG041689) at the University of Pennsylvania, funded by NIA. Harmonized phenotypes were provided by the ADSP Phenotype Harmonization Consortium (ADSP-PHC), funded by NIA (U24 AG074855, U01 AG068057 and R01 AG059716) and Ultrascale Machine Learning to Empower Discovery in Alzheimer’s Disease Biobanks (AI4AD, U01 AG068057). This research was supported in part by the Intramural Research Program of the National Institutes of health, National Library of Medicine. Contributors to the Genetic Analysis Data included Study Investigators on projects that were individually funded by NIA, and other NIH institutes, and by private U.S. organizations, or foreign governmental or nongovernmental organizations.

The ADSP Phenotype Harmonization Consortium (ADSP-PHC) is funded by NIA (U24 AG074855, U01 AG068057 and R01 AG059716). The harmonized cohorts within the ADSP-PHC include:Lthe Anti-Amyloid Treatment in Asymptomatic Alzheimer’s study (A4 Study), a secondary prevention trial in preclinical Alzheimer’s disease, aiming to slow cognitive decline associated with brain amyloid accumulation in clinically normal older individuals. The A4 Study is funded by a public-private-philanthropic partnership, including funding from the National Institutes of Health-National Institute on Aging, Eli Lilly and Company, Alzheimer’s Association, Accelerating Medicines Partnership, GHR Foundation, an anonymous foundation and additional private donors, with in-kind support from Avid and Cogstate. The companion observational Longitudinal Evaluation of Amyloid Risk and Neurodegeneration (LEARN) Study is funded by the Alzheimer’s Association and GHR Foundation. The A4 and LEARN Studies are led by Dr. Reisa Sperling at Brigham and Women’s Hospital, Harvard Medical School and Dr. Paul Aisen at the Alzheimer’s Therapeutic Research Institute (ATRI), University of Southern California. The A4 and LEARN Studies are coordinated by ATRI at the University of Southern California, and the data are made available through the Laboratory for Neuro Imaging at the University of Southern California. The participants screening for the A4 Study provided permission to share their de-identified data in order to advance the quest to find a successful treatment for Alzheimer’s disease. We would like to acknowledge the dedication of all the participants, the site personnel, and all of the partnership team members who continue to make the A4 and LEARN Studies possible. The complete A4 Study Team list is available on: a4study.org/a4-study-team.; the Adult Changes in Thought study (ACT), U01 AG006781, U19 AG066567; Alzheimer’s Disease Neuroimaging Initiative (ADNI): Data collection and sharing for this project was funded by the Alzheimer’s Disease Neuroimaging Initiative (ADNI) (National Institutes of Health Grant U01 AG024904) and DOD ADNI (Department of Defense award number W81XWH-12-2-0012). ADNI is funded by the National Institute on Aging, the National Institute of Biomedical Imaging and Bioengineering, and through generous contributions from the following: AbbVie, Alzheimer’s Association; Alzheimer’s Drug Discovery Foundation; Araclon Biotech; BioClinica, Inc.; Biogen; Bristol-Myers Squibb Company; CereSpir, Inc.; Cogstate; Eisai Inc.; Elan Pharmaceuticals, Inc.; Eli Lilly and Company; EuroImmun; F. Hoffmann-La Roche Ltd and its affiliated company Genentech, Inc.; Fujirebio; GE Healthcare; IXICO Ltd.;Janssen Alzheimer Immunotherapy Research & Development, LLC.; Johnson & Johnson Pharmaceutical Research & Development LLC.; Lumosity; Lundbeck; Merck & Co., Inc.;Meso Scale Diagnostics, LLC.; NeuroRx Research; Neurotrack Technologies; Novartis Pharmaceuticals Corporation; Pfizer Inc.; Piramal Imaging; Servier; Takeda Pharmaceutical Company; and Transition Therapeutics. The Canadian Institutes of Health Research is providing funds to support ADNI clinical sites in Canada. Private sector contributions are facilitated by the Foundation for the National Institutes of Health (www.fnih.org). The grantee organization is the Northern California Institute for Research and Education, and the study is coordinated by the Alzheimer’s Therapeutic Research Institute at the University of Southern California. ADNI data are disseminated by the Laboratory for Neuro Imaging at the University of Southern California; Estudio Familiar de Influencia Genetica en Alzheimer (EFIGA): 5R37AG015473, RF1AG015473, R56AG051876; the Health & Aging Brain Study – Health Disparities (HABS-HD), supported by the National Institute on Aging of the National Institutes of Health under Award Numbers R01AG054073, R01AG058533, R01AG070862, P41EB015922, and U19AG078109; the Korean Brain Aging Study for the Early Diagnosis and Prediction of Alzheimer’s disease (KBASE), which was supported by a grant from Ministry of Science, ICT and Future Planning (Grant No: NRF-2014M3C7A1046042); Memory & Aging Project at Knight Alzheimer’s Disease Research Center (MAP at Knight ADRC): The Memory and Aging Project at the Knight-ADRC (Knight-ADRC). This work was supported by the National Institutes of Health (NIH) grants R01AG064614, R01AG044546, RF1AG053303, RF1AG058501, U01AG058922 and R01AG064877 to Carlos Cruchaga. The recruitment and clinical characterization of research participants at Washington University was supported by NIH grants P30AG066444, P01AG03991, and P01AG026276. Data collection and sharing for this project was supported by NIH grants RF1AG054080, P30AG066462, R01AG064614 and U01AG052410. We thank the contributors who collected samples used in this study, as well as patients and their families, whose help and participation made this work possible. This work was supported by access to equipment made possible by the Hope Center for Neurological Disorders, the Neurogenomics and Informatics Center (NGI: https://neurogenomics.wustl.edu/) and the Departments of Neurology and Psychiatry at Washington University School of Medicine; National Alzheimer’s Coordinating Center (NACC): The NACC database is funded by NIA/NIH Grant U24 AG072122. SCAN is a multi-institutional project that was funded as a U24 grant (AG067418) by the National Institute on Aging in May 2020. Data collected by SCAN and shared by NACC are contributed by the NIA-funded ADRCs as follows: P30 AG062429 (PI James Brewer, MD, PhD), P30 AG066468 (PI Oscar Lopez, MD), P30 AG062421 (PI Bradley Hyman, MD, PhD), P30 AG066509 (PI Thomas Grabowski, MD), P30 AG066514 (PI Mary Sano, PhD), P30 AG066530 (PI Helena Chui, MD), P30 AG066507 (PI Marilyn Albert, PhD), P30 AG066444 (PI John Morris, MD), P30 AG066518 (PI Jeffrey Kaye, MD), P30 AG066512 (PI Thomas Wisniewski, MD), P30 AG066462 (PI Scott Small, MD), P30 AG072979 (PI David Wolk, MD), P30 AG072972 (PI Charles DeCarli, MD), P30 AG072976 (PI Andrew Saykin, PsyD), P30 AG072975 (PI David Bennett, MD), P30 AG072978 (PI Neil Kowall, MD), P30 AG072977 (PI Robert Vassar, PhD), P30 AG066519 (PI Frank LaFerla, PhD), P30 AG062677 (PI Ronald Petersen, MD, PhD), P30 AG079280 (PI Eric Reiman, MD), P30 AG062422 (PI Gil Rabinovici, MD), P30 AG066511 (PI Allan Levey, MD, PhD), P30 AG072946 (PI Linda Van Eldik, PhD), P30 AG062715 (PI Sanjay Asthana, MD, FRCP), P30 AG072973 (PI Russell Swerdlow, MD), P30 AG066506 (PI Todd Golde, MD, PhD), P30 AG066508 (PI Stephen Strittmatter, MD, PhD), P30 AG066515 (PI Victor Henderson, MD, MS), P30 AG072947 (PI Suzanne Craft, PhD), P30 AG072931 (PI Henry Paulson, MD, PhD), P30 AG066546 (PI Sudha Seshadri, MD), P20 AG068024 (PI Erik Roberson, MD, PhD), P20 AG068053 (PI Justin Miller, PhD), P20 AG068077 (PI Gary Rosenberg, MD), P20 AG068082 (PI Angela Jefferson, PhD), P30 AG072958 (PI Heather Whitson, MD), P30 AG072959 (PI James Leverenz, MD); National Institute on Aging Alzheimer’s Disease Family Based Study (NIA-AD FBS): U24 AG056270; Religious Orders Study (ROS): P30AG10161,R01AG15819, R01AG42210; Memory and Aging Project (MAP - Rush): R01AG017917, R01AG42210; Minority Aging Research Study (MARS): R01AG22018, R01AG42210; the Texas Alzheimer’s Research and Care Consortium (TARCC), funded by the Darrell K Royal Texas Alzheimer’s Initiative, directed by the Texas Council on Alzheimer’s Disease and Related Disorders; Washington Heights/Inwood Columbia Aging Project (WHICAP): RF1 AG054023;and Wisconsin Registry for Alzheimer’s Prevention (WRAP): R01AG027161 and R01AG054047. Additional acknowledgments include the National Institute on Aging Genetics of Alzheimer’s Disease Data Storage Site (NIAGADS, U24AG041689) at the University of Pennsylvania, funded by NIA.

## References

1. Alzheimer’s Association. Alzheimer’s disease facts and figures. Alzheimers Dement. 2025;21.

2. Bellenguez C, Küçükali F, Jansen IE, Kleineidam L, Moreno-Grau S, Amin N, et al. New insights into the genetic etiology of Alzheimer’s disease and related dementias. Nat Genet. 2022;54(4):412–36.

3. Medway CW, Abdul-Hay S, Mims T, Ma L, Bisceglio G, Zou F, et al. ApoE variant p.V236E is associated with markedly reduced risk of Alzheimer’s disease. Mol Neurodegener. 2014;9(1):11.

4. He SY, Su WM, Wen XJ, Lu SJ, Cao B, Yan B, et al. Non-genetic risk factors of Alzheimer’s disease: an updated umbrella review. J Prev Alzheimers Dis. 2024;11(4):917–27.

5. Srinivas N, Rachakonda S, Kumar R. Telomeres and telomere length: a general overview. Cancers (Basel). 2020;12(3):558.

6. Demanelis K, Jasmine F, Chen LS, Chernoff M, Tong L, Delgado D, et al. Determinants of telomere length across human tissues. Science. 2020;369(6509):eaaz6876.

7. Broer L, Codd V, Nyholt DR, Deelen J, Mangino M, Willemsen G, et al. Meta-analysis of telomere length in 19,713 subjects reveals high heritability, stronger maternal inheritance, and a paternal age effect. Eur J Hum Genet. 2013;21(10):1163–8.

8. Liu Y, Song YE, Lynn A, Wang W, Miskimen K, Fuzzell SL, et al. Telomere length, aging, and cognitive function in the Midwestern Amish. HGG Adv. 2025;7(1):100533.

9. Bountziouka V, Musicha C, Allara E, Kaptoge S, Wang Q, Di Angelantonio E, et al. Modifiable traits, healthy behaviours, and leukocyte telomere length: a population-based study in UK Biobank. Lancet Healthy Longev. 2022;3(5):e321–31.

10. Lu W, Zhang Y, Liu D, Songyang Z, Wan M. Telomeres—structure, function, and regulation. Exp Cell Res. 2013;319(2):133–41.

11. Gruber HJ, Semeraro MD, Renner W, Herrmann M. Telomeres and age-related diseases. Biomedicines. 2021;9(10):1335.

12. Cai Z, Yan LJ, Ratka A. Telomere shortening and Alzheimer’s disease. Neuromol Med. 2013;15(1):25–48.

13. Burren OS, Dhindsa RS, Deevi SV, Wen S, Nag A, Mitchell J, et al. Genetic architecture of telomere length in 462,666 UK Biobank whole-genome sequences. Nat Genet. 2024;56(9):1832–40.

14. Rodríguez-Fernández B, Vilor-Tejedor N, Arenaza-Urquijo EM, Sánchez-Benavides G, Suárez-Calvet M, Operto G, et al. Genetically predicted telomere length and Alzheimer’s disease endophenotypes: a Mendelian randomization study. Alzheimers Res Ther. 2022;14:167.

15. Nudelman KN, Lin J, Lane KA, Nho K, Kim S, Faber KM, et al. Telomere shortening in the Alzheimer’s disease neuroimaging initiative cohort. J Alzheimers Dis. 2019;71(1):33–43.

16. Kuan XY, Fauzi NS, Ng KY, Bakhtiar A. Exploring the causal relationship between telomere biology and Alzheimer’s disease. Mol Neurobiol. 2023;60(8):4169–83.

17. Fani L, Hilal S, Sedaghat S, Broer L, Licher S, Arp PP, et al. Telomere length and the risk of Alzheimer’s disease: the Rotterdam study. J Alzheimers Dis. 2020;73(2):707–14.

18. Hansen ME, Hunt SC, Stone RC, Horvath K, Herbig U, Ranciaro A, et al. Shorter telomere length in Europeans than in Africans due to polygenetic adaptation. Hum Mol Genet. 2016;25(11):2324–30.

19. Greenfest-Allen E, Valladares O, Kuksa PP, Gangadharan P, Lee WP, Cifello J, et al. NIAGADS Alzheimer’s GenomicsDB: a resource for exploring Alzheimer’s disease genetic and genomic knowledge. Alzheimers Dement. 2024;20(2):1123–36.

20. Naj AC, Lin H, Vardarajan BN, White S, Lancour D, Ma Y, et al. Quality control and integration of genotypes from two calling pipelines for whole genome sequence data in the Alzheimer’s Disease Sequencing Project. Genomics. 2019;111(4):808–18.

21. Lee WP, Choi SH, Shea MG, Cheng PL, Dombroski BA, Pitsillides AN, et al. Association of common and rare variants with Alzheimer’s disease in more than 13,000 diverse individuals with whole-genome sequencing from the Alzheimer’s Disease Sequencing Project. Alzheimers Dement. 2024;20(12):8470–83.

22. Leung YY, Valladares O, Chou YF, Lin HJ, Kuzma AB, Cantwell L, et al. VCPA: genomic variant calling pipeline and data management tool for Alzheimer’s Disease Sequencing Project. Bioinformatics. 2019;35(10):1768–70.

23. Jin H, Chien S, Meijer E, et al. Learning from clinical consensus diagnosis in India to facilitate automatic classification of dementia: machine learning study. JMIR Ment Health. 2021;8:e27113.

24. Gogarten SM, Sofer T, Chen H, Yu C, Brody JA, Thornton TA, et al. Genetic association testing using the GENESIS R/Bioconductor package. Bioinformatics. 2019;35(24):5346–8.

25. Purcell S, Neale B, Todd-Brown K, Thomas L, Ferreira MAR, Bender D, et al. PLINK: a toolset for whole-genome association and population-based linkage analysis. Am J Hum Genet. 2007;81:559–75.

26. Reynolds D. Gaussian mixture models. In: Li SZ, Jain AK, editors. Encyclopedia of Biometrics. Springer; 2015. p. 827–32.

27. Ding Z, Mangino M, Aviv A, UK10K Consortium, Spector T, Durbin R. Estimating telomere length from whole genome sequence data. Nucleic Acids Res. 2014;42(9):e75.

28. Mazidi M, Penson P, Banach M. Association between telomere length and complete blood count in US adults. Arch Med Sci. 2017;13:601–5.

29. Choi SW, O’Reilly PF. PRSice-2: Polygenic Risk Score software for biobank-scale data. Gigascience. 2019;8(7):giz082.

30. Chen MH, Raffield LM, Mousas A, Sakaue S, Huffman JE, Moscati A, et al. Trans-ethnic and ancestry-specific blood-cell genetics in 746,667 individuals from five global populations. Cell. 2020;182(5):1198–213.

31. R Core Team. R: A language and environment for statistical computing. Vienna: R Foundation for Statistical Computing; 2021.

32. Han B, Eskin E. Random-effects model aimed at discovering associations in meta-analysis of genome-wide association studies. Am J Hum Genet. 2011;88(5):586–98.

33. Cingolani P, Platts A, Wang LL, Coon M, Nguyen T, Wang L, et al. A program for annotating and predicting the effects of single nucleotide polymorphisms, SnpEff: SNPs in the genome of Drosophila melanogaster strain w1118; iso-2; iso-3. Fly (Austin). 2012;6(2):80–92.

34. McLaren W, Gil L, Hunt SE, Riat HS, Ritchie GR, Thormann A, et al. The Ensembl Variant Effect Predictor. Genome Biol. 2016;17:122.

35. Lewis MJ, Wang S. locuszoomr: an R package for visualizing publication-ready regional gene locus plots. Bioinformatics Adv. 2025;5(1):vbaf006.

36. Mostafavi S, Gaiteri C, Sullivan SE, White CC, Tasaki S, Xu J, et al. A molecular network of the aging human brain provides insights into the pathology and cognitive decline of Alzheimer’s disease. Nat Neurosci. 2018;21(6):811–9.

37. De Jager PL, Ma Y, McCabe C, Xu J, Vardarajan BN, Felsky D, et al. A multi-omic atlas of the human frontal cortex for aging and Alzheimer’s disease research. Sci Data. 2018;5:180142.

38. Love MI, Huber W, Anders S. Moderated estimation of fold change and dispersion for RNA-seq data with DESeq2. Genome Biol. 2014;15(12):550.

39. Hackenhaar FS, Josefsson M, Adolfsson AN, Landfors M, Kauppi K, Hultdin M, et al. Short leukocyte telomeres predict 25-year Alzheimer’s disease incidence in non-APOE ε4-carriers. Alzheimers Res Ther. 2021;13:130.

40. Wikgren M, Karlsson T, Nilbrink T, Nordfjäll K, Hultdin J, Sleegers K, et al. APOE ε4 is associated with longer telomeres, and longer telomeres among ε4 carriers predict worse episodic memory. Neurobiol Aging. 2012;33(2):335–44.

41. Lansdorp PM. Sex differences in telomere length, lifespan, and embryonic dyskerin levels. Aging Cell. 2022;21(5):e13614.

42. Perez-Branguli F, Zagar Y, Shanley DK, Graef IA, Chédotal A, Mitchell KJ. Reverse signaling by semaphorin-6A regulates cellular aggregation and neuronal morphology. PLoS One. 2016;11(7):e0158686.

43. Rünker AE, Little GE, Suto F, Fujisawa H, Mitchell KJ. Semaphorin-6A controls guidance of corticospinal tract axons at multiple choice points. Neural Dev. 2008;3:34.

44. Zhang L, Qi Z, Li J, Li M, Du X, Wang S, et al. Roles and mechanisms of axon-guidance molecules in Alzheimer’s disease. Mol Neurobiol. 2021;58(7):3290–307.

45. Yuan J, Huang R, Nao J, Dong X. The role of Semaphorin 3A in the pathogenesis and progression of Alzheimer’s disease and other aging-related diseases: a comprehensive review. Pharmacol Res. 2025;11:107732.

46. Carulli D, De Winter F, Verhaagen J. Semaphorins in adult nervous system plasticity and disease. Front Synaptic Neurosci. 2021;13:672891.

47. Hatanaka Y, Kawasaki T, Abe T, Shioi G, Kohno T, Hattori M, et al. Semaphorin-6A–Plexin A2/A4 interactions with radial glia regulate migration termination of superficial layer cortical neurons. iScience. 2019;21:359–74.

48. Chung S, Yang J, Kim HJ, Hwang EM, Lee W, Suh K, et al. Plexin-A4 mediates amyloid-β–induced tau pathology in Alzheimer’s disease animal model. Prog Neurobiol. 2021;203:102075.

49. Jun G, Asai H, Zeldich E, Drapeau E, Chen C, Chung J, et al. PLXNA4 is associated with Alzheimer’s disease and modulates tau phosphorylation. Ann Neurol. 2014;76(3):379–92.

50. Dieck S, Sanmartí-Vila L, Langnaese K, Richter K, Kindler S, Soyke A, et al. Bassoon, a novel zinc-finger CAG/glutamine-repeat protein selectively localized at the active zone of presynaptic nerve terminals. J Cell Biol. 1998;142(2):499–509.

51. Kaladiyil AP, Kukkle PL. Bassoon, presynaptic scaffolding protein: narrative review in health and disease. Eur J Neurosci. 2025;61(5):e70066.

52. Martínez P, Patel H, You Y, Jury N, Perkins A, Lee-Gosselin A, et al. Bassoon contributes to tau-seed propagation and neurotoxicity. Nat Neurosci. 2022;25:1597–607.

53. Schubach M, Maass T, Nazaretyan L, et al. CADD v1.7: using protein language models, regulatory CNNs, and other nucleotide-level scores to improve genome-wide variant predictions. Nucleic Acids Res. 2024;52:D1143–54.

54. Dey A, Allen JN, Fraser JW, Snyder LM, Tian Y, Zhang L, et al. Neuroprotective role of the RON receptor tyrosine kinase underlying central nervous system inflammation in health and disease. Front Immunol. 2018;9:513.

55. Adamu A, Li S, Gao F, Xue G. The role of neuroinflammation in neurodegenerative diseases: current understanding and future therapeutic targets. Front Aging Neurosci. 2024;16:1347987.

56. Karati D, Kumar D. Tyrosine kinase as therapeutic target of neurodegenerative disorders. Brain Disord. 2025;100:193.

57. Li J, Chanda D, Shiri-Sverdlov R, Neumann D. MSP: an emerging player in metabolic syndrome. Cytokine Growth Factor Rev. 2015;26(1):75–82.

58. Ezkurdia-Lasarte A, Ramírez-Gil M, Solas-Zubiaurre M. Metabolic syndrome as a risk factor for Alzheimer’s disease: a focus on insulin resistance. Front Aging Neurosci. 2025;In press.

59. Song B, Davis K, Liu XS, Lee HG, Smith M, Liu X. Inhibition of polo-like kinase 1 reduces β-amyloid-induced neuronal cell death in Alzheimer’s disease. Aging (Albany NY). 2011;3(9):846–57.

60. Chen LL, Wang YB, Song JX, Deng WK, Lu JH, Ma LL, et al. Phosphoproteome-based kinase activity profiling reveals the critical role of MAP2K2 and PLK1 in neuronal autophagy. Autophagy. 2017;13(11):1969–80.

61. von Zglinicki T. Oxidative stress shortens telomeres. Trends Biochem Sci. 2002;27(7):339–44.

62. Zhang J, Rane G, Dai X, Shanmugam MK, Arfuso F, Samy RP, et al. Ageing and the telomere connection: an intimate relationship with inflammation. Ageing Res Rev. 2016;25:55–69.

63. Kinney JW, Bemiller SM, Murtishaw AS, Leisgang AM, Salazar AM, Lamb BT. Inflammation as a central mechanism in Alzheimer’s disease. Alzheimers Dement (Transl Res Clin Interv). 2018;4:575–90.

64. Heneka MT, Carson MJ, El Khoury J, Landreth GE, Brosseron F, Feinstein DL, et al. Neuroinflammation in Alzheimer’s disease. Lancet Neurol. 2015;14(4):388–405.

65. Li Z, Shue F, Zhao N, Shinohara M, Bu G. APOE2: protective mechanism and therapeutic implications for Alzheimer’s disease. Mol Neurodegener. 2020;15:63.

66. Schächter F, Faure-Delanef L, Guénot F, Rouger H, Froguel P, Lesueur-Ginot L, et al. Genetic associations with human longevity at the APOE and ACE loci. Nat Genet. 1994;6(1):29–32.

67. Mahoney ER, Dumitrescu L, Seto M, Nudelman KN, Buckley RF, Gifford KA, et al. Telomere length associations with cognition depend on Alzheimer’s disease biomarkers. Alzheimers Dement. 2019;5:883–90.

68. Needham BL, Salerno S, Roberts E, Boss J, Allgood KL, Mukherjee B. Do Black/White differences in telomere length depend on socioeconomic status? Biodemography Soc Biol. 2020;65(4):287–312.

