## Supplemental Figures for "Interactive Effects of Telomere Length and Genetic Variants on Alzheimer Disease Risk Across Multiple Ancestral Populations"

**Additional file 1**

**Fig. S1.** Workflow of the sample level Quality Control (QC)

**Fig. S2.** Principle component (PC) analysis showing population clusters based on first and second PCs in the total dataset

**Fig. S3.** Telomere density distribution across total and different ancestries

**Fig. S4.** Telomere length distribution in short and long groups by ancestry

**Fig. S5.** GWAS results in the European ancestry dataset

**Fig. S6.** GWAS results in the Caribbean Hispanic dataset

**Fig. S7.** GWAS results in the Native American Hispanic dataset

**Fig. S8.** GWAS results in the African American dataset

**Fig. S9.** Locus zoom plots for the top-ranked SNPs

**Fig. S1.** Workflow of the sample level Quality Control (QC). WGS = Whole Genome Sequencing; AD = Alzheimer Disease.

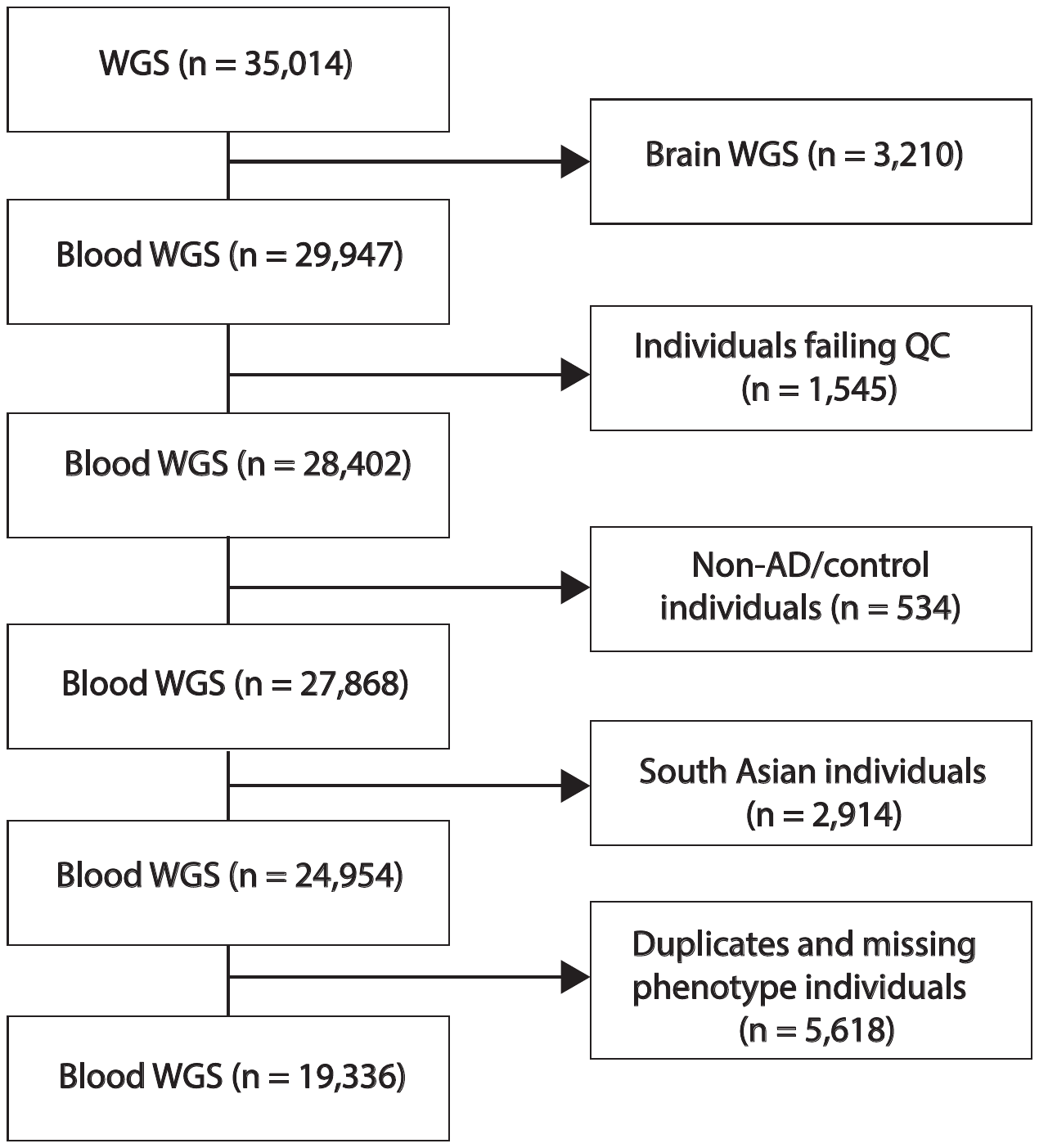

**Fig. S2.** Principle component (PC) analysis showing population clusters based on first and second PCs in the total dataset. AA = African American; EA = European ancestry; CH = Caribbean Hispanic; NAH = Native American Hispanic.

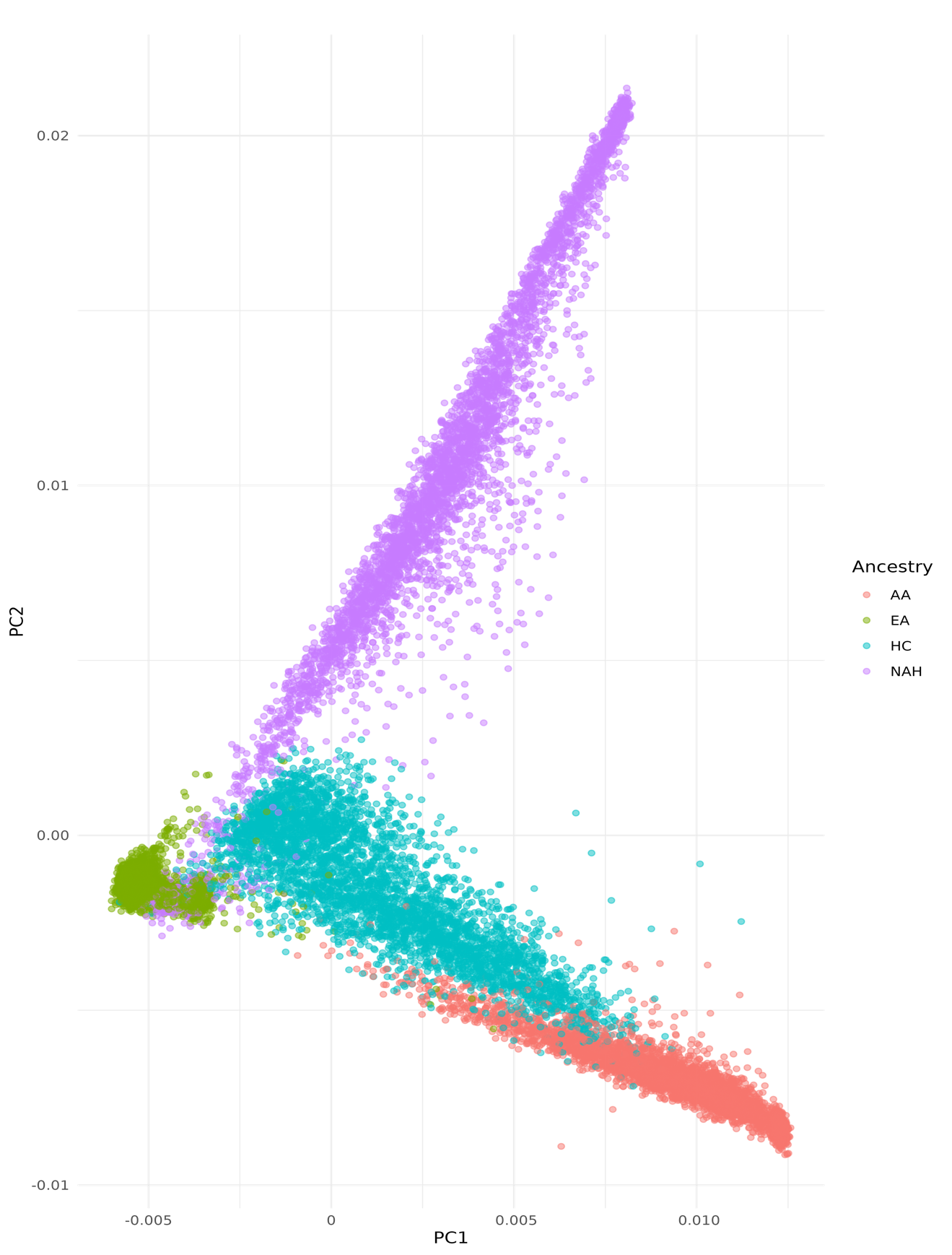

**Fig. S3.** Telomere length density distribution by ancestry

.

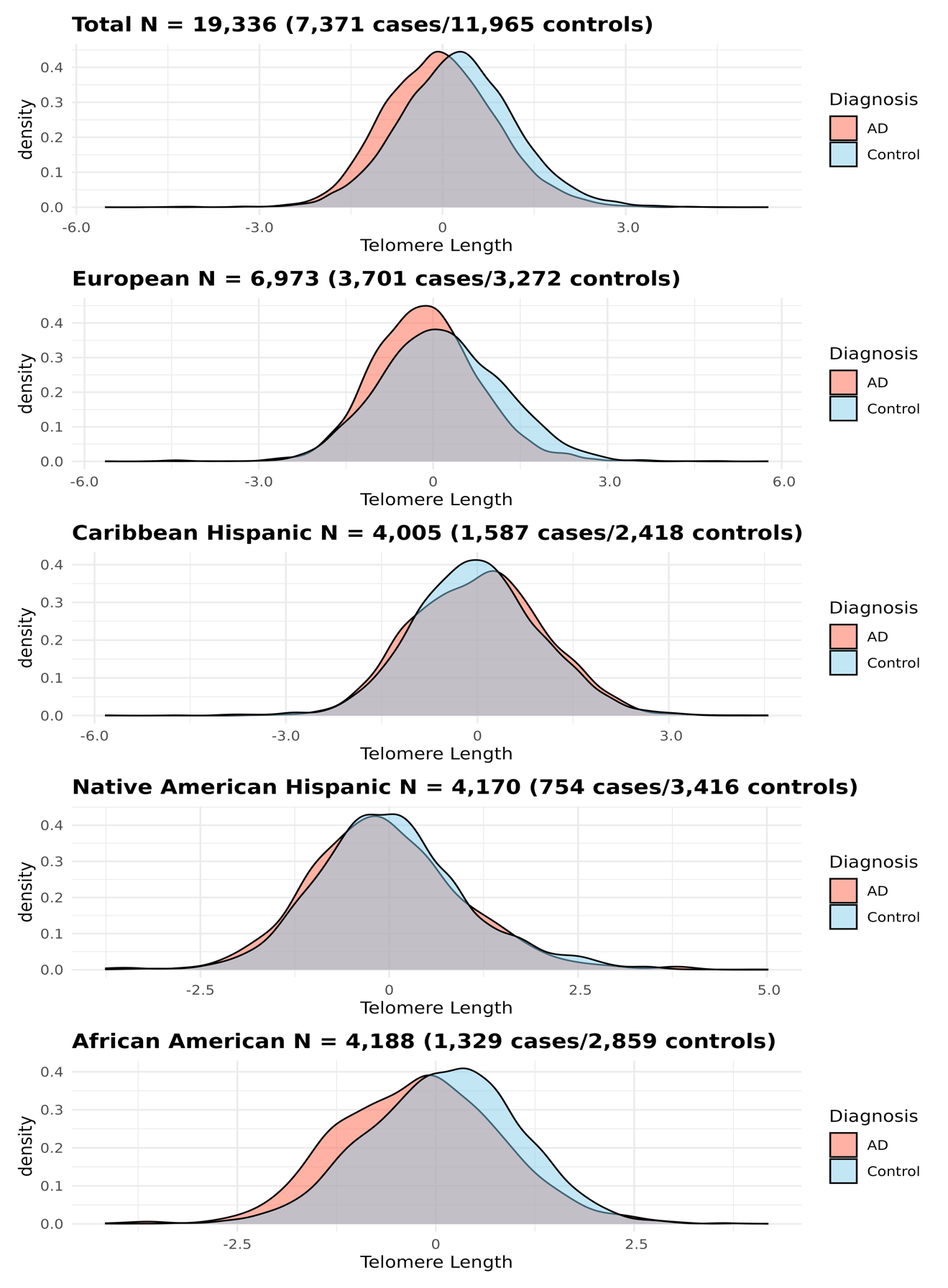

**Fig. S4.** Telomere length distribution in short and long groups by ancestry.

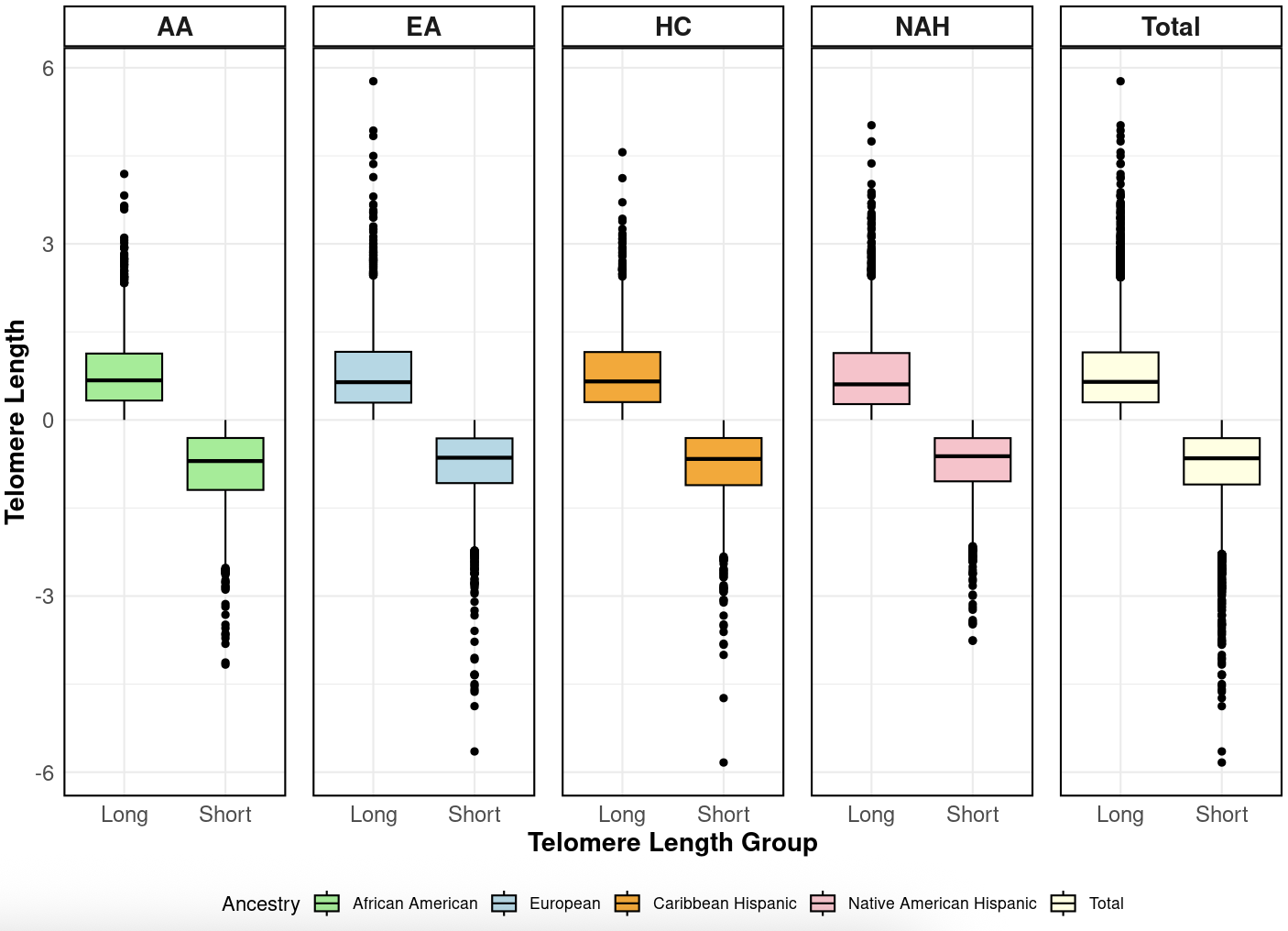

**Fig. S5.** GWAS results in the European ancestry dataset. **A.** QQ plot showing measure of genomic inflation (lambda). **B.** Manhattan plot showing significant TL x SNP interactions with AD. Red and blue horizontal lines indicate thresholds for study-wide (*P*<5.00x10^-8^) and suggestive (*P*<1.00x10^-5^) significance, respectively.

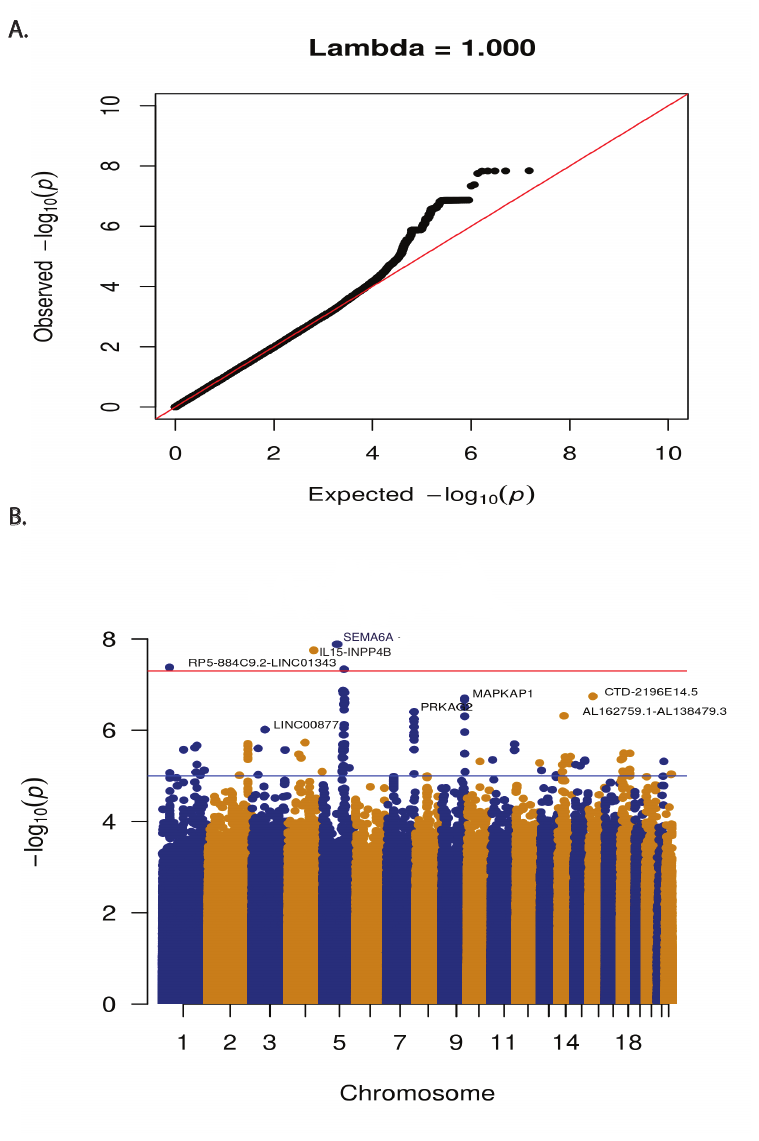

**Fig. S6.** GWAS results in the Caribbean Hispanic ancestry dataset. **A.** QQ plot showing measure of genomic inflation (lambda). **B.** Manhattan plot showing significant TL x SNP interactions with AD. Red and blue horizontal lines indicate thresholds for study-wide (*P*<5.00x10^-8^) and suggestive (*P*<1.00x10^-5^) significance, respectively.

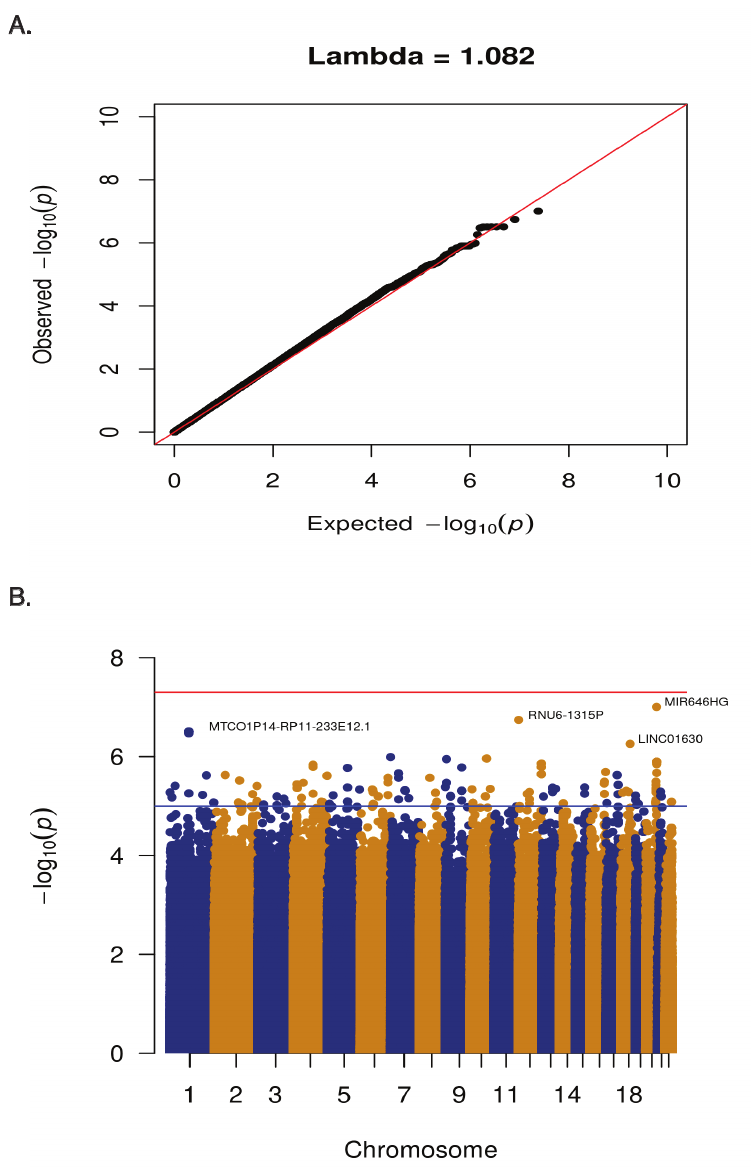

**Fig. S7.** GWAS results in the Native American Hispanic ancestry dataset. **A.** QQ plot showing measure of genomic inflation (lambda). **B.** Manhattan plot showing significant TL x SNP interactions with AD. Red and blue horizontal lines indicate thresholds for study-wide (*P*<5.00x10^-8^) and suggestive (*P*<1.00x10^-5^) significance, respectively.

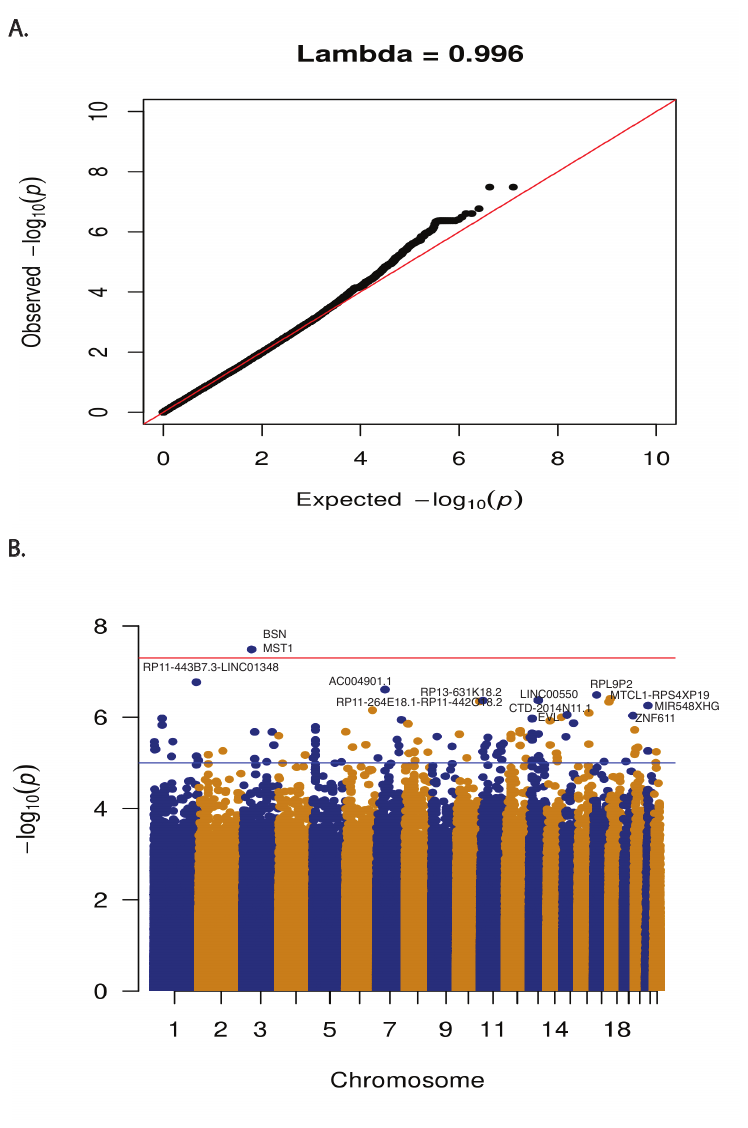

**Fig. S8.** GWAS results in the African American ancestry dataset. **A.** QQ plot showing measure of genomic inflation (lambda). **B.** Manhattan plot showing significant TL x SNP interactions with AD. Red and blue horizontal lines indicate thresholds for study-wide (*P*<5.00x10^-8^) and suggestive (*P*<1.00x10^-5^) significance, respectively.

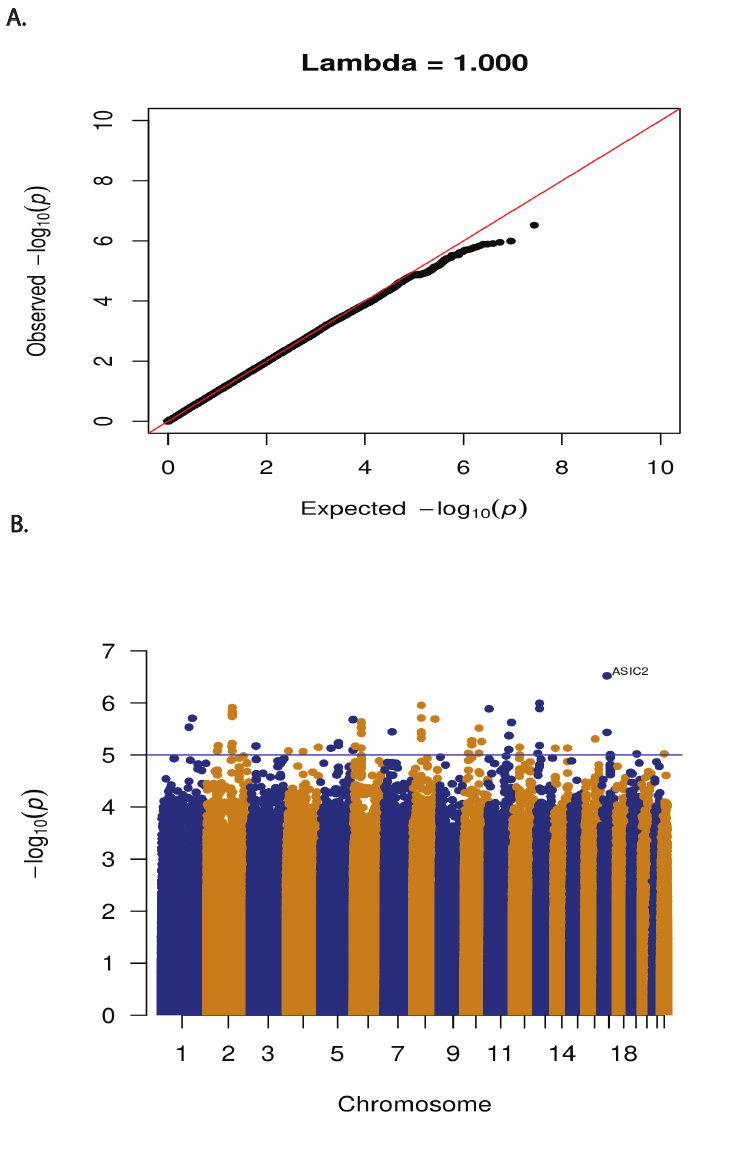

**Fig. S9.** Locus zoom plots for the top-ranked SNPs. SNPs are color-coded indicating their correlation with the top rank SNP (purple diamond).

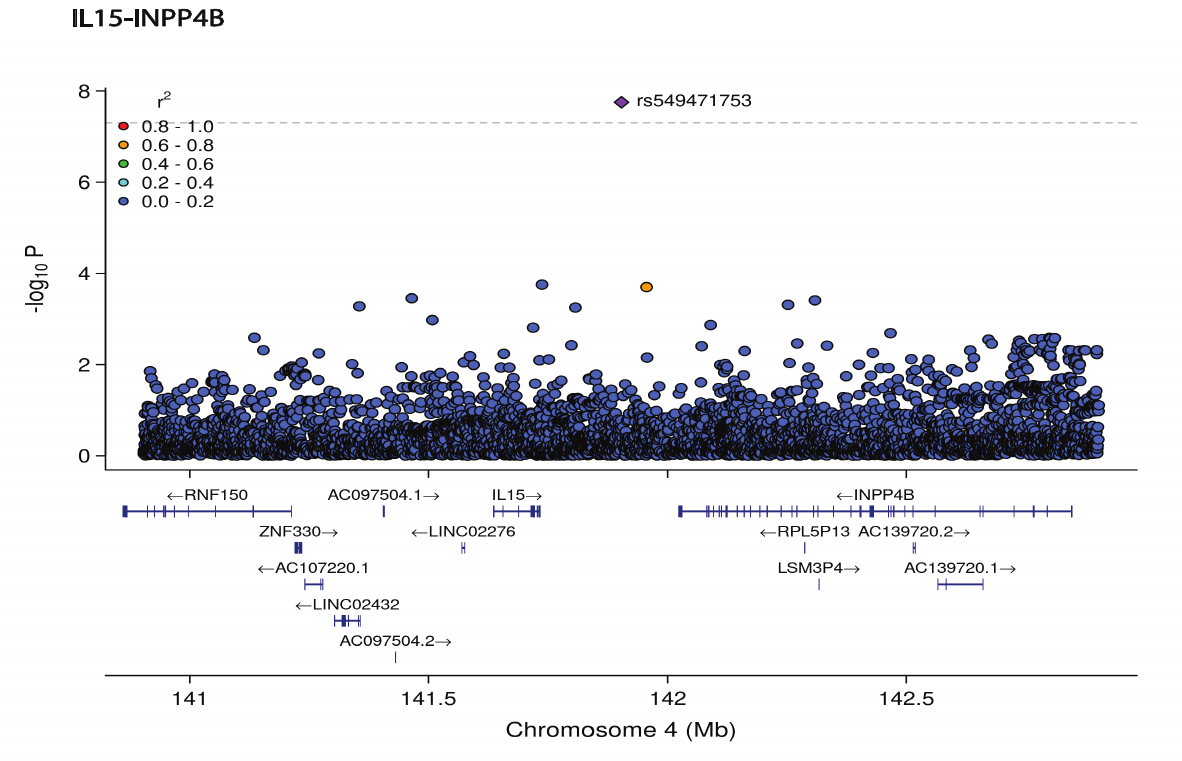

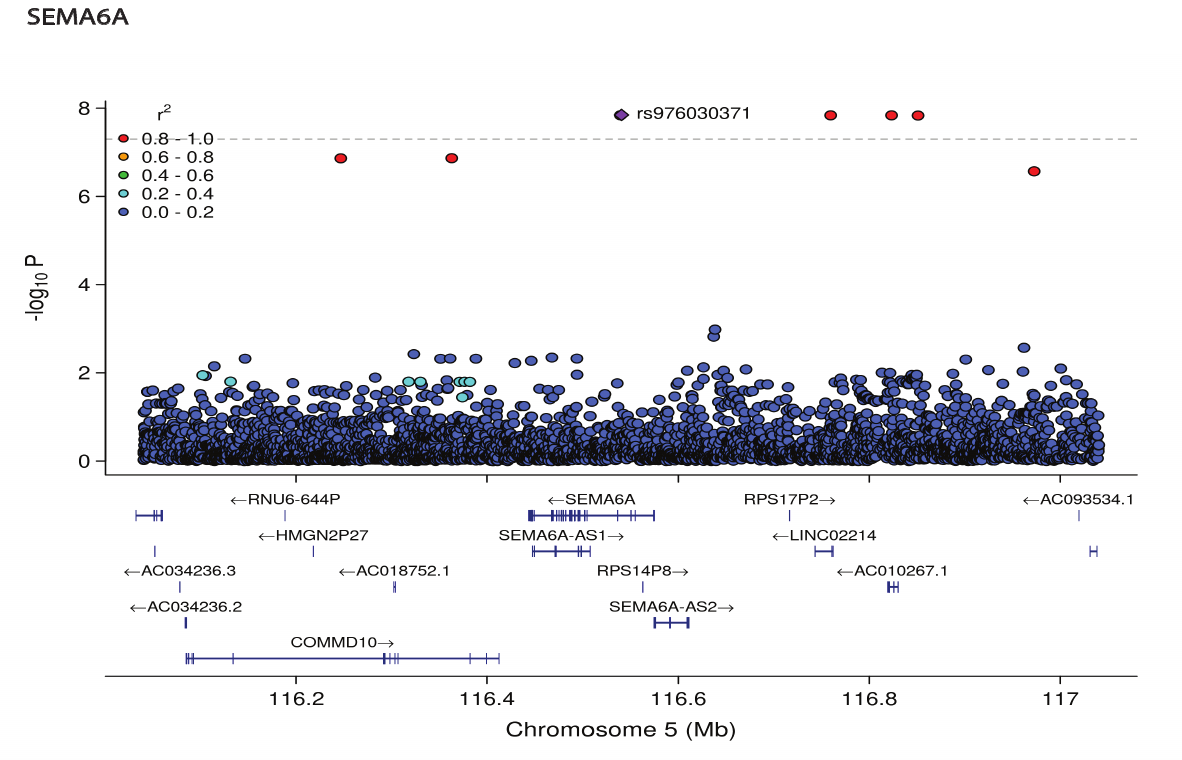

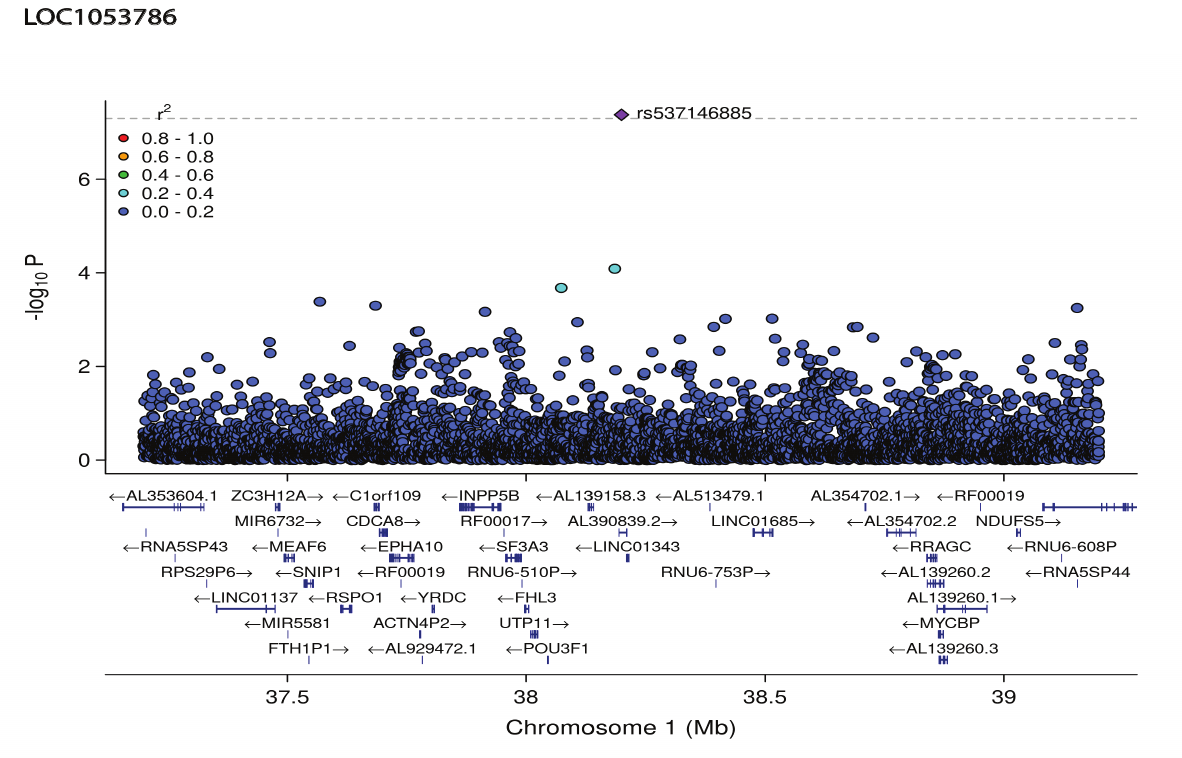

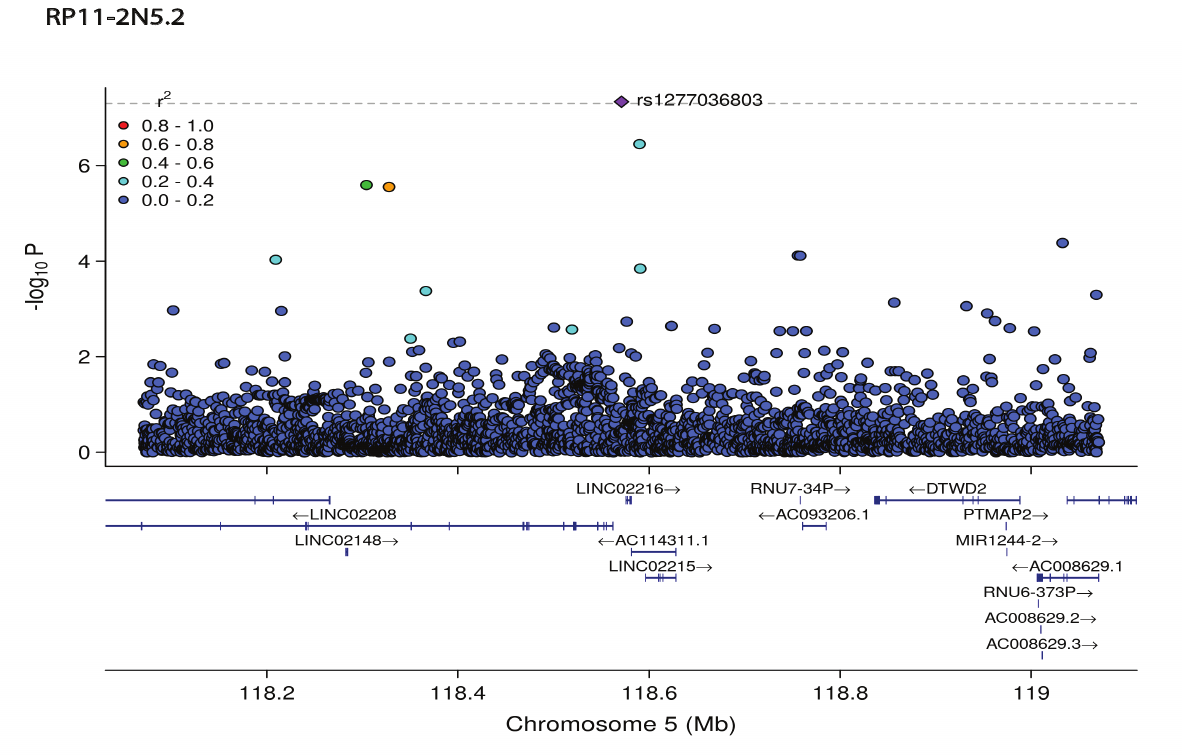

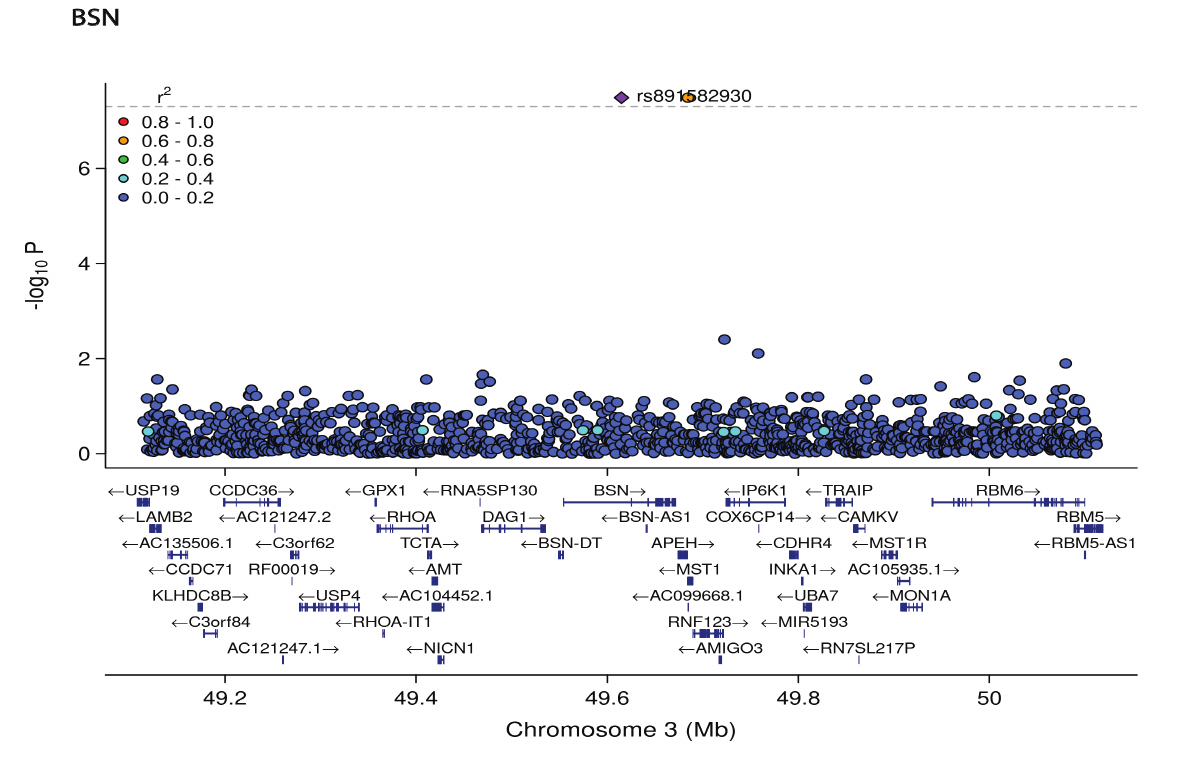
